# Effectiveness, Explainability and Reliability of Machine Meta-Learning Methods for Predicting Mortality in Patients with COVID-19: Results of the Brazilian COVID-19 Registry

**DOI:** 10.1101/2021.11.01.21265527

**Authors:** Bruno Barbosa Miranda de Paiva, Polianna Delfino-Pereira, Claudio Moisés Valiense de Andrade, Virginia Mara Reis Gomes, Maria Clara Pontello Barbosa Lima, Maira Viana Rego Souza-Silva, Marcelo Carneiro, Karina Paula Medeiros Prado Martins, Thaís Lorenna Souza Sales, Rafael Lima Rodrigues de Carvalho, Magda C. Pires, Lucas Emanuel F. Ramos, Rafael T. Silva, Adriana Falangola Benjamin Bezerra, Alexandre Vargas Schwarzbold, Aline Gabrielle Sousa Nunes, Amanda de Oliveira Maurílio, Ana Luiza Bahia Alves Scotton, André Soares de Moura Costa, Andriele Abreu Castro, Bárbara Lopes Farace, Christiane Corrêa Rodrigues Cimini, Cíntia Alcantara De Carvalho, Daniel Vitório Silveira, Daniela Ponce, Elayne Crestani Pereira, Euler Roberto Fernandes Manenti, Evelin Paola de Almeida Cenci, Fernanda Barbosa Lucas, Fernanda D’Athayde Rodrigues, Fernando Anschau, Fernando Antonio Botoni, Fernando Graça Aranha, Frederico Bartolazzi, Gisele Alsina Nader Bastos, Giovanna Grunewald Vietta, Guilherme Fagundes Nascimento, Helena Carolina Noal, Helena Duani, Heloisa Reniers Vianna, Henrique Cerqueira Guimarães, Isabela Moraes Gomes, Jamille Hemétrio Salles Martins Costa, Jéssica Rayane Corrêa Silva da Fonseca, Júlia Di Sabatino Santos Guimarães, Júlia Drumond Parreiras de Morais, Juliana Machado Rugolo, Joanna D’arc Lyra Batista, Joice Coutinho de Alvarenga, José Miguel Chatkin, Karen Brasil Ruschel, Leila Beltrami Moreira, Leonardo Seixas de Oliveira, Liege Barella Zandoná, Lílian Santos Pinheiro, Luanna da Silva Monteiro, Lucas de Deus Sousa, Luciane Kopittke, Luciano de Souza Viana, Luis César de Castro, Luisa Argolo Assis, Luisa Elem Almeid Santos, Máderson Alvares de Souza Cabral, Magda Cesar Raposo, Maiara Anschau Floriani, Maria Angélica Pires Ferreira, Maria Aparecida Camargos Bicalho, Mariana Frizzo de Godoy, Matheus Carvalho Alves Nogueira, Meire Pereira de Figueiredo, Milton Henriques Guimarães-Júnior, Mônica Aparecida de Paula De Sordi, Natália da Cunha Severino Sampaio, Neimy Ramos de Oliveira, Pedro Ledic Assaf, Raquel Lutkmeier, Reginaldo Aparecido Valacio, Renan Goulart Finger, Roberta Senger, Rochele Mosmann Menezes, Rufino de Freitas Silva, Saionara Cristina Francisco, Silvana Mangeon Mereilles Guimarães, Silvia Ferreira Araújo, Talita Fischer Oliveira, Tatiana Kurtz, Tatiani Oliveira Fereguetti, Thainara Conceição de Oliveira, Thulio Henrique Oliveira Diniz, Yara Cristina Neves Marques Barbosa Ribeiro, Yuri Carlotto Ramires, Marcos André Gonçalves, Milena Soriano Marcolino

## Abstract

**Objective:** To provide a thorough comparative study among state-of-the-art machine learning methods and statistical methods for determining in-hospital mortality in COVID-19 patients using data upon hospital admission; to study the reliability of the predictions of the most effective methods by correlating the probability of the outcome and the accuracy of the methods; to investigate how explainable are the predictions produced by the most effective methods.

**Materials and Methods:** De-identified data were obtained from COVID-19 positive patients in 36 participating hospitals, from March 1 to September 30, 2020. Demographic, comorbidity, clinical presentation and laboratory data were used as training data to develop COVID-19 mortality prediction models. Multiple machine learning and traditional statistics models were trained on this prediction task using a folded cross-validation procedure, from which we assessed performance and interpretability metrics.

**Results:** The Stacking of machine learning models improved over the previous state-of-the-art results by more than 26% in predicting the class of interest (death), achieving 87.1% of AUROC and macro F1 of 73.9%. We also show that some machine learning models can be very interpretable and reliable, yielding more accurate predictions while providing a good explanation for the ‘why’.

**Conclusion:** The best results were obtained using the meta-learning ensemble model – Stacking. State-of the art explainability techniques such as SHAP-values can be used to draw useful insights into the patterns learned by machine-learning algorithms. Machine-learning models can be more explainable than traditional statistics models while also yielding highly reliable predictions.

## Introduction

The number of patients with coronavirus disease 2019 (COVID-19), as well as the related deaths, have increased exponentially since the World Health Organization declared it a pandemic on March 2020. Up to September 24, 2021, there are over 230 million cumulative cases and 4.7 million deaths reported worldwide (1). Although over 6 billion doses of COVID-19 vaccines have been administered worldwide, due to an uneven and slow rollout, variants are emerging and outbreaks continue especially in poorer countries, meaning that COVID-19 will be an issue governments worldwide will need to keep grappling with (1,2).

Given the current scenario, there is an urgent need for an early disease stratification tool upon hospital admission, to allow the early identification of risk of death in patients with COVID-19, assisting in the management of disease and optimising resource allocation, hopefully assisting to save lives during the pandemic. Although several scores have been proposed for the early assessment of COVID-19 patients at hospital admission, the majority of them are bounded by methodological flaws and technological limitations, meaning that reliable prognostic prediction models are scarce (3–5).

A state-of-the-art method for this prediction task has recently been proposed by our group with the development of a new risk score - ABC_2_-SPH - using traditional statistical methods (least absolute shrinkage and selection operator - LASSO regression), which exploits a rich set of information, including patient’s demographics, comorbidities, vital signs and laboratory parameters at the time of presentation, for assessing prognosis in COVID-19 patients. The model has shown high discriminatory value (AUROC 0.844, 95% CI 0.829 to 0.859), confirmed in the Brazilian (0.859 [95% CI 0.833 to 0.885]) and Spanish (0.899 [95% CI 0.864 to 0.934]) validation cohorts, and with better discrimination ability than other existing scores (4).

In this context, artificial intelligence (AI), and more specifically machine learning (ML), techniques have been explored in various fields for dealing with the pandemic, such as detecting outbreaks, diagnosis, interpretation of chest imaging exams to detect COVID-19 lung disease, vaccines development and prognosis prediction (6,7), but comprehensive comparative studies to investigate whether ML techniques have superior performance when compared to models using traditional statistical methods are still scarce.

Indeed in several other contexts (8) ML techniques have demonstrated superior effectiveness (i.e., accuracy) when compared to traditional statistical methods (e.g., logistic regression), due for instance, their capability of dealing with collinearity and redundancy, as well the ability to find non-linear correlations among the variables. However, current studies in the mortality prediction for COVID-19 using ML techniques are limited, regarding either methodological or technological aspects.

In this scenario, the contributions of this article are fivefold. First, we provide a **thorough comparative study** among state-of-the-art ML methods, including many modern techniques, such as transformer and convolutional neural networks, boosting algorithms, support vector machines (SVM), k-nearest neighbors, as well as state-of-the-art statistical methods, represented by ABC_2_-SPH, in the task of determining **in-hospital mortality** in COVID-19 patients using data **upon hospital admission**.

Second, given the profusion and diversity of the compared methods, we investigate the effectiveness of meta-learning ensemble strategies, most notably Stacking (9), that combine the methods’ outputs (probabilities), in order to exploit the ML methods’ strengths and overcome their limitations.

Third, we study the **reliability** of the predictions of the most effective methods by correlating the probability of the outcome and the effectiveness (accuracy) of the methods. Few studies have investigated this important aspect of the predictions, which has practical impact in the applicability of the methods. Fourth, we investigated how **interpretable** (or explainable) are the predictions produced by the most effective methods using modern interpretability tools. Explainability is an essential aspect of the task if ML methods are to be trusted and actually used by practitioners.

Finally, we provide a discussion on the adequacy of AUROC as an evaluation metric for highly imbalanced and skewed datasets commonly found in health-related problems, as is the case of our COVID-19 study.

## Related Work

This study also included a narrative review of the scientific literature on existing prediction models for COVID-19 mortality using artificial intelligence techniques. These models were identified through a literature search of Medline and MedRxiv, with no language or date restrictions, using the search not indexed terms: “COVID-19”, “SARS-CoV-2”, combined with “mortality”, “prognosis”, “risk factors”, “hospitalizations” or “score”. The last search was performed in August 2021.

Following the narrative analysis, our initial search highlighted papers that satisfied our search criteria, removing duplicates, leaving relevant articles for the title and abstract review. Text screening retained 76 studies included in the S1 Table.

The existing literature largely focuses on American and Chinese hospitals, represented together by 53.94% of studies. In fact, models validated in one country cannot be extrapolated to the population as a whole, since there is heterogeneity among countries in different characteristics such as populations features (including genetics, race, ethnicity, prevalence of comorbidities), socioeconomic factors, access to healthcare, and the healthcare systems themselves (hospitals patient load, practice and available resources) (10).

Another important point is the sample size. Larger population studies are needed to allow certain metrics of model performance to be estimated with more accurate and reliable results. In contrast, smaller samples reduce the ability to identify risk factors and increase the likelihood of overfitting (11). Among the analyzed models, 17.10% were developed and validated with a modest sample of 500-1000 patients, and 35.52% used even a smaller sample, with less than 500 patients. Only 47.36% of the studies used a sample with more than 1000 patients. Our sample used in this study has 5032 patients.

Most of the studies (60.52%) used traditional statistical methods, including multivariate logistic regression, LASSO and Cox regression analysis. Artificial intelligence techniques were used in 39.47% of studies, among them stands out machine learning, including random forest (RF), XGBoost and SVM. And only a very small percentage (11.8%) of works exploit modern neural network methods in their studies as we do in ours.

Overall, the majority of developed models are limited by methodological bias, with for example, absence of external validation in 51.31%, so the assessment of accuracy in those studies may be overestimated. A minority of them (around 23.68%) reported having followed the methodological recommendations from Transparent Reporting of a multivariable prediction model for Individual Prognosis Or Diagnosis (TRIPOD) (11).

The model performance was evaluated in most studies, by area under the curve (AUC), and the mean AUC for training ranged from 0.64-0.96 for traditional statistical methods, and from 0.74-0.96 for models using AI techniques. However, due to the very high skewness of the datasets (i.e., mortality corresponds to a very low percentage of the cases in the datasets, in other words, the non-death class dominates the distribution) neither AUC nor accuracy are adequate metrics (12).

To properly assess the performance of different models, it is of utmost importance to use other metrics that consider imbalance issues, such as macro-average F1-score (macro-F1), used in 13.33% studies. For example, Li et al (2020) developed a deep-learning model and a risk-score system based on 55 clinical variables and observed that the most crucial biomarkers distinguishing patients at mortality imminent risk, were age, lactate dehydrogenase, procalcitonin, cardiac troponin, C-reactive protein and oxygen saturation (13). The deep-learning model predicted mortality with an AUC of 0.852 and 0.844, for training and testing, respectively, which is considered excellent (13). However, the performance of the proposed algorithm on training and testing datasets measured by the F1-score was 0.642 and 0.616, respectively (13).

Finally, few studies (S1 Table) deep analyzed the impact of the variables in the final model or on the final model outcome. Additionally, most studies do not investigate how reliable the made predictions are in terms of the correlation between the probability of the prediction and the accuracy. This analysis has implications on the practical use of this technology. An accurate but unreliable method has its practical applicability diminished. We explicitly tackle these issues in our study.

## Materials and Methods

This is a substudy of the Brazilian COVID-19 Registry, a multi-hospital cohort study previously described (14). Complying with the study protocol, adult patients with laboratory-confirmed COVID-19 according to the World Health Organization criteria, admitted consecutively in any of the 36 participating hospitals, from March 1 to September 30, 2020 were enrolled. Individuals were not included if they were transferred between hospitals and data from the first or last hospitals was not available, as well those who were admitted for other reasons and developed COVID-19 symptoms during their stay (4).

Trained hospital staff or interns collected demographic information, clinical characteristics, laboratory and outcome data from medical records. A prespecified case report form was used, applying Research Electronic Data Capture (REDCap) tools (15). To ensure data quality, comprehensive data quality checks were undertaken. Error checking code was developed in R to identify data entry errors, as previously described (4), and the results were sent to each center for checking and correction before further data analysis.

Variables used to develop the models were obtained upon hospital presentation. A set of potential predictor features for in-hospital mortality was selected a priori, as recommended, from demographics, home medications, past medical history, clinical features, and laboratory values (S2 Table) (4,11). Laboratory exams were performed at the discretion of the treating physician. The ABC_2_-SPH score is composed by age, blood urea nitrogen, number of comorbidities, C-reactive protein, SpO_2_/FiO_2_ ratio, platelet count and heart rate (4).

### Data analysis

The development, validation and reporting of the models followed guidance from the TRIPOD (Transparent Reporting of a multivariable prediction model for Individual Prognosis Or Diagnosis) checklist and the Prediction model Risk of Bias Assessment Tool (PROBAST) (11,16). All data was fully anonymized. At that time 36 Brazilian hospitals participated in the cohort, located in 17 cities, from five Brazilian states (4).

A total of 5032 patients were admitted between March 1, 2020 and September 31, 2020, and the full group was used to perform a 10-fold cross validation procedure, which was repeated 3 times (at a total of 30 performance measurements for each of the classifiers presented in our study). The overall study population included 45.9% women, with a mean age of 60 (standard deviation [SD] 17) years, 1367 (27.17%) needed mechanical ventilation and 1014 (20.15%) died.

In order to properly assess the performance of different models, we chose to use three different metrics, each assessed through the aforementioned 10-fold cross validation procedure, for each classifier. Our evaluation metrics include both micro-average and macro-average F1-score (micro-F1 and macro-F1), and the area (AUROC) under the receiver operating curve (ROC-Curve). While more common in healthcare-related literature, the AUROC values can be misleading, especially when there is significant class imbalance (17), and even more so when the class of interest is the rare one (which is usually the case). Therefore, we included the micro and macro-F1 scores as evaluation metrics. The F1 score is the harmonic mean between precision and recall scores, for each class (i.e. one score to estimate how well the model can predict which patients will die, and one to estimate the same regarding which patients will not die). The “average” part, described as either “micro” or “macro”, refers to how these results are aggregated. In “macro” averaging, all classes are taken as equally important, while in “micro” averaging, class imbalance is not accounted for in the final result and all individual predictions are considered equally important (18).

As for the specific models compared in our study, we trained two modern neural network benchmarks -- the FNet transformer, with and without virtual adversarial training, which is a regularization technique -- and a deep convolutional Resnet. We also experimented with a support vector machine classifier, a boosting model (microsoft research’s Light Gradient Boosting Machine), and the K-nearest neighbors algorithm, as well as a stacking of these methods.

We compare these ML alternatives to traditional statistical methods, including a Generalized Additive Model (GAM), which has rarely been used in this scenario before, and LASSO regression, the current state-of-the-art. GAM was used before in ABC_2_-SPH, but only to select variables for the lasso regression, which yielded an inferior result when compared to LASSO regression, whereas in our work, we directly tune GAM to the classification task, thus obtaining better results, as we shall see.

The choice of neural networks to include in our study was motivated by current state-of-the-art methods, even though, in general, neural networks tend to perform better in situations where massive amounts of data are available, which is not our case, as we have a relatively small data sample (12,19). Usually, the ability to compare distant input positions in the query vectors is related to the neural network’s depth. Transformer architectures, as introduced by Vaswani et al (2017), gained rapid success due to their capacity of doing so in a constant number of operations, achieving state-of-the-art results in many tasks (20). That is the reason we chose a FNet Transformer classifier. For comparison purposes, we also included a Resnet model, which held similar success for image classification, due to the capacity of building very deep networks. Due to the relative drop in performance of neural networks when fewer data samples are present in training, we also include a training variant where we perform virtual adversarial training, as introduced in Miyato et al (2017), in which the model’s decision boundary is smoothed in the most anisotropic direction through a gradient-based approximation (21).

Additionally, we included a standard support vector machine classifier, which learns a separation hyperplane between classes, while maximising the separation margin, and a K-nearest neighbors classifier, which yields predictions based on spatial similarities between training samples and new query points. Motivated by the results shown in Shwartz et al (2021) (22), we included a boosting algorithm (LightGBM), which is usually an effective model in tabular data, as concluded in Ke et al (2017) (23). As the final classifier, we included a meta-learning ensemble-based Stacking model, which learns to combine the prediction outputs of all previous classifiers in order to improve classification effectiveness. We compare these methods to Generalised Additive Models (GAM) and LASSO regression, the latter being the current state-of-the-art model for this task, as demonstrated in our previous work.

We ran all classification tests using a 10 fold cross validation procedure, after which we calculated confidence intervals for each result, and confirmed statistical significance by applying a Wilcoxon signed-rank test with 95% confidence.

For the parametrization of our models, we used the values presented in Table 1, where the values in brackets are evaluated in the validation set of the cross validation process. For deep network models we use the early stop to optimize the model, which optimizes the weights until the model has no significant improvement in the validation set.

**Table 1.** Parameterization of methods.

| Method | Parametrization |
| --- | --- |
| SVM | C: [$10^{-3}$, $10^{-2}$, $10^{-1}$, $10^0$, $10^1$, $10^2$]<br>Kernel: [linear, rbf, poly, sigmoid]<br>class_weight: [None, 'balanced'] |
| RF | N-estimators: [10, 50, 100, 200, 500, 1000, 2000] |
| KNN | Neighbors: [2, 4, 8, 16, 32] |
| LASSO | Alpha: [$10^{-3}$, $10^{-2}$, $10^{-1}$, $10^0$, $10^1$, $10^2$] |
| LIGHT_GBM | N-estimators: [10, 50, 100, 200, 500, 1000, 2000]<br>learning_rate: [$10^{-3}$, $10^{-2}$, $10^{-1}$, $30^{-1}$]<br>colsample_by_tree: [0.5, 1.0] |
| CNN | Early Stop |
| FNet | Early Stop |
| FNet + VAT | Early Stop |
| GAM | No tuning |
| Stacking | Meta-Classifer: Logistic Regression, Alpha: [$10^2$] |

List of model names: CNN = convolutional neural network, FNet = fourier transformation neural network, FNet + VAT = fourier transformation neural network with virtual adversarial training, GAM = generalized
additive models, KNN = K-nearest neighbors, LASSO = lasso regression, LIGHT\_GBM = light gradient boosting machines, RF = random forest, SVM = support vector machines, STACKING = a stacking classifier, which combines all others.

## Results and Discussion

Classification results for the prediction of death can be found in Table 1. Neural network models (CNN - convolutional neural networks - and FNet - Fourrier transform neural network - / FNet + VAT - Fourrier transform neural network + virtual adversarial training) produced the worst results while boosting (‘LightGBM’ - Light Gradient Boosting Machine), Stacking and one traditional statistical models (‘Generalized Additive Models - GAM’) produced the best overall results, when considering both, micro and macro-F1, and AUROC. It is interesting to notice that GAM surpassed LASSO, which was used in our ABC_2_-SPH score and was considered the previous state-of-the-art.

The less effective results of the Neural network are somewhat expected as the size of the dataset is not that huge, with fewer than 10 thousand samples. Typically, we expect neural networks of large capacity (millions to billions of parameters) to excel in tasks where very large datasets are available (millions to billions of training instances), which is very rare in health-related problems. In such large-scale datasets, neural networks can capture very complex relationships. However, in smaller sample sizes, they show a remarkable tendency to overfit, hence obtaining poor results in terms of validation error (12,19).

In general, tree-based ensemble models such as random and boosting forests tend to be more robust to small sample sizes and to overfitting, which is exactly the behavior we observed in our experiments (24). SVM and K-nearest neighbors (KNN), which are simpler models, with fewer parameters, also tend to perform reasonably well on smaller datasets being better than the neural network models.

We should stress that the statistical models LASSO regression and mainly Generalized additive models (GAM) showed very competitive results for this data sample. Unexpectedly, GAM was the runner up method considering all metrics, being even better than LASSO and some traditional ML methods such as SVM and KNN. This result contrasts with the one in ABC_2_-SPH, where GAM was used simply to select variables for the LASSO regression. In our work, we directly tuned GAM to the classification task, using the cross-validation procedure, which yielded superior performance. GAM and LightGBM are statistically tied regarding all evaluation metrics considering a Wilcoxon signed-rank test with 95% confidence.

In any case, the best single overall model, with statistical significance, under all considered metrics, was the Stacking model, which is a combination of the output of all other individual models, which, in turn, exploited all the provided features (S2 Table), including demographic data, comorbidities, lifestyle habits, clinical assessment and laboratory data upon hospital admission: age; days from symptom onset; heart and respiratory rate, mechanical ventilation, oxygen inspiration fraction, platelets, urea, C-reactive protein, lactate, gasometry results (pH, pO_2_, pCO_2_, bicarbonate), hemogram parameters (hemoglobin, neutrophils, lymphocytes, neutrophils to lymphocytes ratio, platelets) and sodium upon hospital admission. When considering micro and macro-F1, F1 for death and AUROC at the task of predicting death, Stacking was significantly (statistically) better than all other models. The largest gains were in F1 to predict death with gains of up more than 26% over LASSO, the previous state-of-the-art.

Indeed, we observe in Table 2 that the combination of models by means of Stacking yields statistically significant improvements over all the best individual single models (RF, Boosting and GAM), allowing us to better discriminate between patients with higher clinical risk at admission time. The Stacking technique improves the F1-score results for the class of interest (death) by 7% over RF, by 5% for LightGBM and by 6% for GAM, which were the three individual best models in this metric. The combination of models based on different classification premises, potentially made stacking more robust. If a single classifier makes a wrong prediction, the others can still make corrections (since the predictions are independent), increasing the robustness of the final stacking model.

**Table 2.** Micro-F1, macro-F1 and AUROC results for the prediction of COVID-19 in-hospital death.

|  | **MICRO-F1** | **MACRO-F1** | **F1 (DEATH)** | **F1 (NO DEATH)** | **AUROC** |
| --- | --- | --- | --- | --- | --- |

|  | mean | CI | mean | CI | mean | CI | mean | CI | mean | CI |
| --- | --- | --- | --- | --- | --- | --- | --- | --- | --- | --- |
| **KNN** | 0.807 | 0.002 | 0.492 | 0.007 | 0.091 | 0.014 | 0.892 | 0.001 | 0.781 | 0.010 |
| **FNet + VAT** | 0.810 | 0.013 | 0.677 | 0.020 | 0.470 | 0.038 | 0.884 | 0.009 | 0.772 | 0.019 |
| **FNet** | 0.814 | 0.008 | 0.686 | 0.017 | 0.486 | 0.030 | 0.887 | 0.005 | 0.789 | 0.015 |
| **CNN** | 0.815 | 0.013 | 0.693 | 0.016 | 0.500 | 0.026 | 0.886 | 0.009 | 0.796 | 0.016 |
| **SVM** | 0.839 | 0.010 | 0.691 | 0.031 | 0.478 | 0.058 | 0.904 | 0.005 | 0.833 | 0.012 |
| **LASSO** | 0.842 | 0.009 | 0.677 | 0.024 | 0.446 | 0.044 | 0.908 | 0.005 | 0.859 | 0.006 |
| **LIGHT_GBM** | 0.846 | 0.008 | 0.723 | 0.016 | 0.538 | 0.028 | 0.908 | 0.005 | 0.865 | 0.008 |
| **GAM** | 0.847 | 0.006 | 0.720 | 0.014 | 0.532 | 0.026 | 0.908 | 0.003 | 0.855 | 0.012 |
| **RF** | 0.850 | 0.005 | 0.717 | 0.013 | 0.524 | 0.024 | 0.911 | 0.003 | 0.863 | 0.007 |
| **STACKING** | **0.855** | 0.007 | **0.739** | 0.018 | **0.564** | 0.032 | **0.913** | 0.004 | **0.871** | 0.007 |

List of model names, from top to bottom (ordered by MicF1): CNN = convolutional neural network, FNet = fourier transformation neural network, FNet + VAT = fourier transformation neural network with virtual adversarial training, GAM = generalized additive models, KNN = K-nearest neighbors, LASSO = lasso regression, LIGHT\_GBM = light gradient boosting machines, RF = random forest, SVM = support vector machines, STACKING = a stacking classifier, which combines all others.

As an additional final analysis, given the popularity of this metric in the health domain, we generated ROC curves for all evaluated models, shown in Fig 1. From this Figure, we can see the separation of two distinct groups. There is a group of models with inferior results, composed of neural network models and K-nearest neighbors, and a group of models with superior (indistinguishable) results, consisting of SVM, RF, LightGBM, GAM and the Stacking of models. Despite similarities in the curves and at AUROC values, these classifiers can yield quite different results when compared with micro-F1 and macro-F1, or class-specific F1 scores, which shows that (1) AUROC score is not an adequate metric for evaluating and comparing models, especially in face of high imbalance/skewness and that (2) even though some models, like Stacking and GAM have very similar AUROC scores, their capacity to discriminate relevant outcomes like death is quite different (0.532 F1 score for GAM end 0.564 for Stacking, a significant difference of 6%).

**Fig 1.**
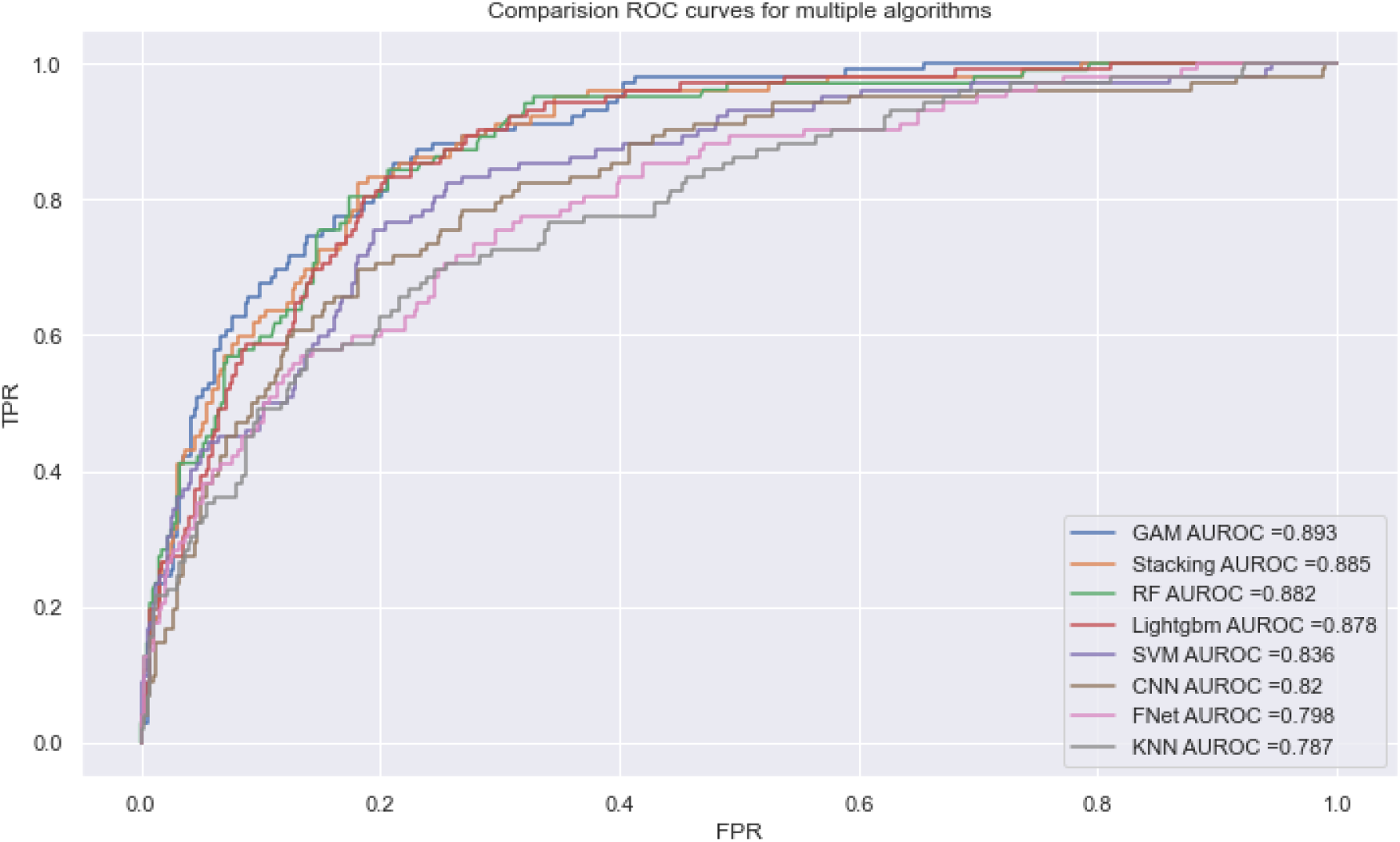
Receiver Operating Characteristic (ROC) Curve comparing multiple models, trained on the prediction of the death outcome.

Another interesting remark is that, using such curves, we can sensibly calibrate the trade-off between sensitivity and specificity, further customizing the way such models can be used. In particular, when applying Stacking, our model can be tailored to the early identification of high-risk patients with good discrimination capacity.

### Explainability

Various prognostic factors have been proposed in the stratification of COVID-19 patients, based on their risk of death, that includes clinical, laboratory and radiological variables. Among these risk factors, stand out advanced age, multiple comorbidities on admission (such as hypertension, diabetes mellitus, cardiovascular diseases and others), abnormal levels of C-reactive protein (CRP), lymphocytes, neutrophils, D-dimer, blood urea nitrogen (BUN) and lactate dehydrogenase (LDH).

A very interesting feature of some ML models, in particular decision trees, RF and boosting forests, is the explainability of these models. This is still a very active research area, but modern advances in tools and visualization alternatives allow us to represent which features were most important to the model and at which polarities and intervals. The best model in our tests was the Stacking. However, this is a meta-model whose inputs are the outputs of other classifiers. Because of that, and since we want to explain a classifier that works on the level of the features themselves instead of a meta-level of other classifier outputs, we will provide explanations for the second best model, LightGBM. Furthermore, tree-based boosting and bagging algorithms rank as some of the most explainable machine learning models, and also lead many benchmarks, particularly for tabular data where data samples are not that large. Their unique combination of explainability, reliability and performance, added to the fact that stacking is a meta-classifier are why we will exploit the boosting model (which, in our case, outperformed the bagging model - random forests/RF) to analyse the found correlations among variables.

In a sense, some traditional models, such as regression models, also have a good explainability, as we can assess the coefficients of each attribute to measure how important a feature is. These models however do not measure up to modern tree-based algorithms in many scenarios, especially in cases with larger datasets (25). Another key difference between these models is that, in the case of regression models, we have to explicitly remove collinear variables, but these variables, even though they might not improve classification performance, still yield valid model explanations. In addition to that, tree based models can return explanations in the form of intervals, such as the behavior seen in Fig 2 for sodium and bicarbonate levels, which imply there is a ‘safe interval’ at which death risk is lower, while either extreme (i.e. too low or too high) has a predictive value for the possibility of a COVID-19 related death.

**Fig 2.**
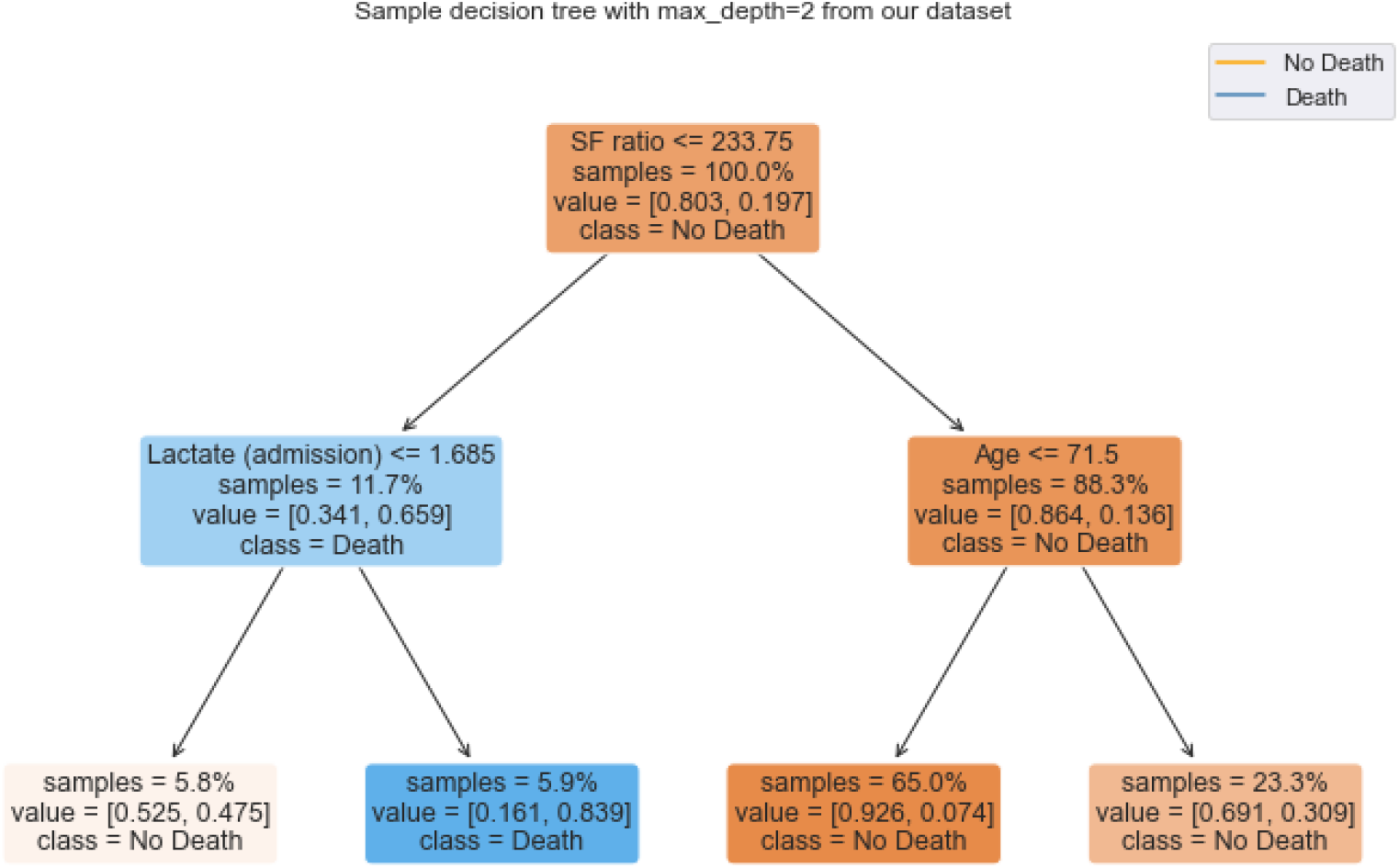
A sample decision tree with depth 2, trained on our dataset. At each level but the last, the first line of text in each box shows the variable and its cut before the split.

From a clinical perspective, our results, shown in the Figures, are in line with a recent study with patients from two hospitals in London, which has shown that hypernatremia and hyponatremia during COVID-19 hospitalization are associated with a higher risk of death and respiratory failure, respectively (26). With regards to bicarbonate, low levels are related to acidosis, and high levels are usually related to advanced chronic obstructive pulmonary disease (COPD) with retention of carbon dioxide, both of them conditions well-known to be associated with worse prognosis in clinical practice (27–29). This sort of non-linearity cannot be captured by simple regression models, since we can only measure how large coefficient values are, and correlate that to the importance of each feature.

In decision tree based algorithms, however, each node represents a feature. The closer to the root (i.e. the ‘first’ node of each tree), the more the feature is able to differentiate the data classes. For example, in Fig 2, feature ‘SF ratio’’ with the value less than 233 and the feature ‘lactate’ with a value less than 1.68 mmol/L results in a subset with 5.9% of the dataset where the ‘death’ outcome is more common.

These algorithms look for the values of the features that further separate the classes, while trying to decrease the coefficient or entropy values of the class label (which are measures of purity and information) in each partition in each decision tree -- this coefficient is called the GINI Index. Such index and the entropy score tend to isolate records that represent the most frequent class in a branch.

In Fig 3, we present SHapley Additive exPlanation (SHAP) values for our boosting model. This is a special type of explainability technique, which allows us to not only probe which features were important to the model, but also which polarities or intervals push predictions to each of the training classes and, additionally, allows us to evaluate why the model predicted any single instance (30).

**Fig 3.**
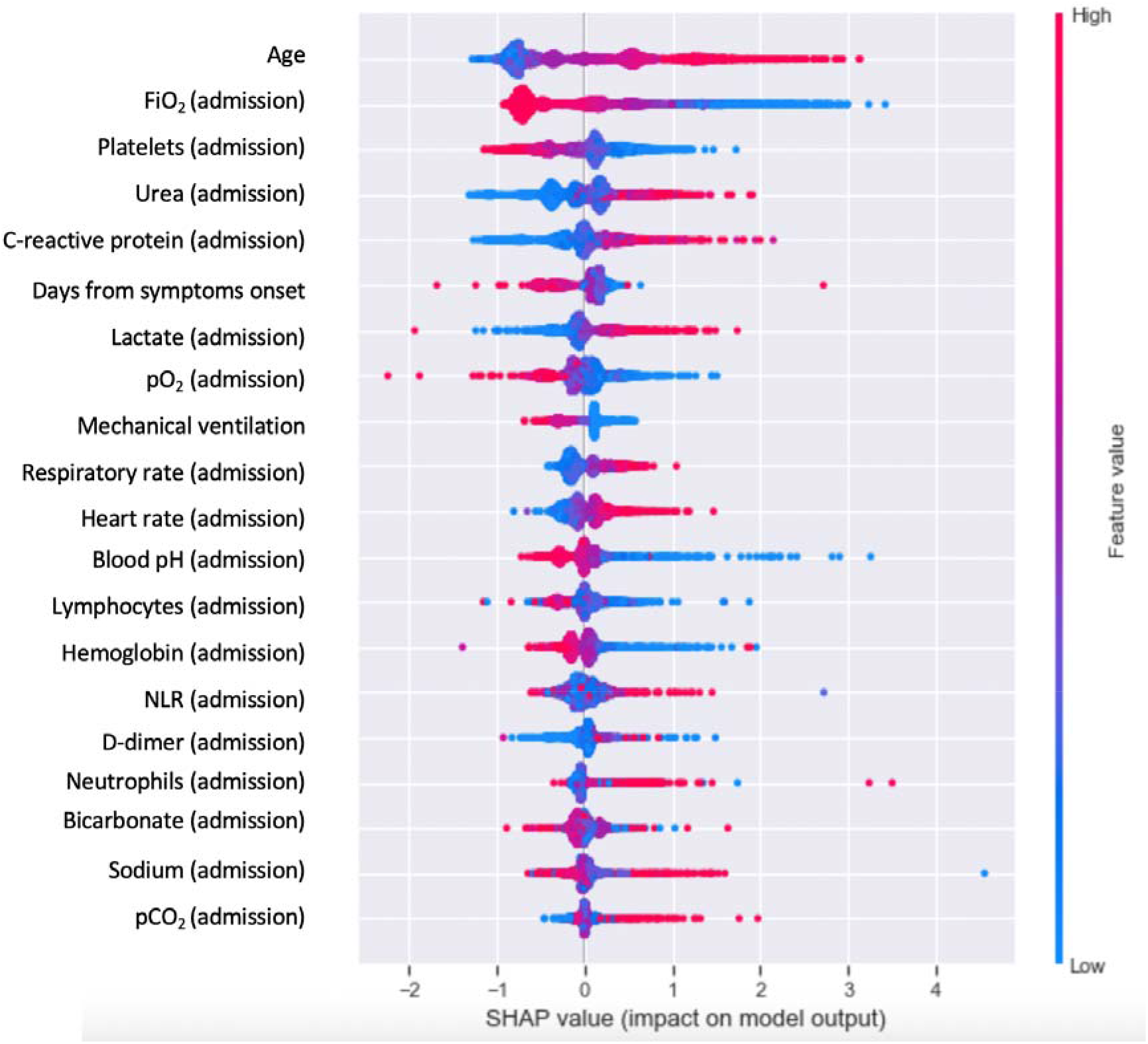
SHAP values for the LightGBM model trained on the prediction of the death outcome.

For any simple model, such as regression, the model itself is a reasonable explanation of what was learned. However, for more complex models, which in turn are capable of learning more complex solutions (provided enough data is present at training time), we cannot use the model to explain itself, since it is a complex solution. In these situations, shapley values build upon the idea that the explanation of a model can itself be a model. This technique was introduced recently by Lundberg et al (2020), and further expands on the explainability of machine learning models, making them even more useful, as they become more interpretable (30).

With the help of SHAP values in Figs 3 and 4, we can extract interesting knowledge from our boosting model, the best individual ML model that works with the base features. We can see for instance that the most important feature in the prediction of death COVID-19 is age. This is coherent with previous medical literature, and serves as an additional validation to the model. Other scores and a recent meta-analysis have shown age as a key prognostic determinant in COVID-19 (31–34). The meta-analysis included more than half million of COVID-19 patients from different countries, and observed that the risk increased exponentially after the fifth decade of life. It is important to highlight that this fact could be influenced by both the physiological aging process and, especially by the individuals functional status and reserve, what may hinder the intrinsic capacity to fight against infections, increasing susceptibility to the infection and severe clinical manifestations (35).

**Fig 4.**
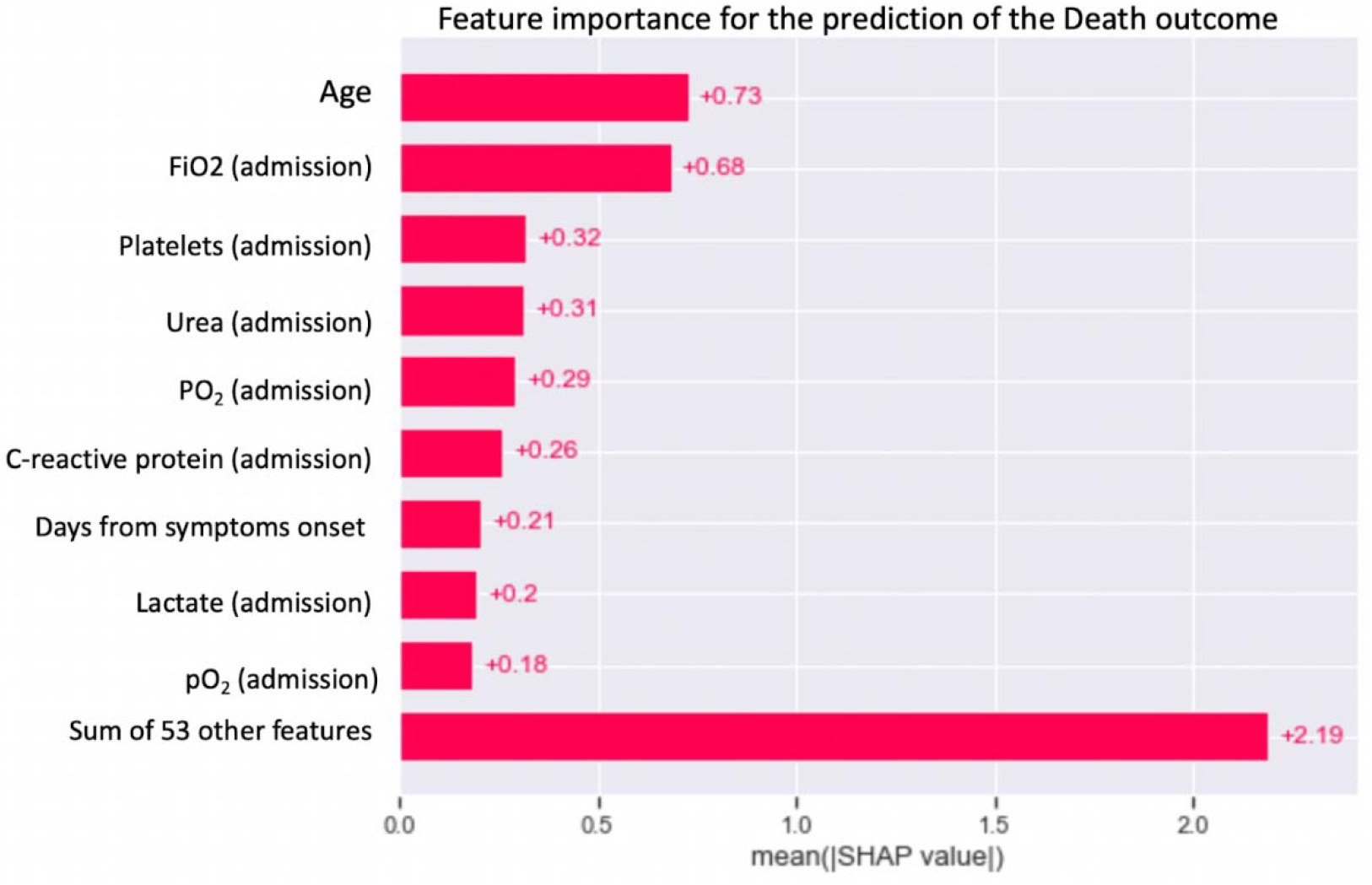
Mean SHAP values for each feature in the prediction of either death.

A recent Brazilian study in a center not included in the present analysis observed that frailty assessed using the Clinical Frailty Scale is a key predictor of COVID-19 prognosis. The authors identified different mortality risks within age and acute morbidity groups. As our study was based on chart review only, we could not assess frailty, but we agree with study authors that it must not be neglected when assessing COVID-19 prognosis. In addition to helping identify patients with a higher risk of death, it can be valuable in guiding evidence-based discussions on realistic goals patients can achieve (35).

The second most important feature is the supplemental oxygen requirement, which, as per Fig 3, lower values (blue tones) indicate higher risk. Although COVID-19 is a multisystem disease, it is well known that lung involvement is the mainstain for assessing disease severity, and oxygen requirement upon hospital admission has been shown to be an independent predictor for severe COVID-19 in several studies (36,37).

Still in this analysis, lower values of platelets also increase risk of mortality, as well as higher levels of urea and C-reactive protein, which was in line with what was previously observed (38). Other studies suggest that C-reactive protein was a marker of a cytokine storm developing in patients with COVID-19 and was associated with the disease mortality (39–41).

An interesting behavior that we can observe with SHAP values and which might not be possible to analyze with a simple regression model, is the one seen in features like admission sodium and bicarbonate serum levels, in which there is a “safe zone” for which risk is lower, but values either too high or too low yield higher risk. This is an intrinsic limitation of regression models, and the variable may be seen as non-significant due to the fact that it is a non-linear association.

As previously mentioned, an important limitation of regression models is collinearity. When exploiting LASSO regression in our previous work (4), we had to exclude some features which had shown to be important in the boosting model due to high collinearity. This may explain the difference in the features included in both models, despite the fact that all features included in both had previous evidence of association with COVID-19 prognosis.

Another interesting remark is shown in Fig 4, in which we can see the relative importance of each feature. Here, again, age is the most important single feature (due to higher mean SHAP value), which is in line with previous studies (3,31,32). In an American study in intensive care units, age has shown higher discriminatory capacity when used in isolation (AUC 0.66) than the Sequential Organ Failure Assessment (SOFA) score (0.55) for mortality prediction, in a cohort study of adult patients from 18 ICUs in the US, with COVID-19 pneumonia. This score is widely used at emergency departments and ICUs worldwide to determine the extent of a person’s organ function or rate of failure (42). In the present study, the remaining features, when combined, yield higher predictive value in this task than just age.

### Reliability

Finally, we investigate issues related to the reliability of the models. Neural network models are, for instance, known for having irregular error rates, regardless of prediction confidence. At the other end of the spectrum, boosting and bagging models tend to have a very interesting reliability profile, with a tendency to have lower error rates at high confidence scores, and higher error rates at lower confidence scores. This generates a very useful perspective, in which we can tune the trade-off between accuracy and sensitivity for some specific classifiers.

Accordingly, we show in Fig 5 the reliability profile for our best model (Stacking). In this Figure, the x-axis shows prediction ranges for the model’s confidence score, while the y-axis shows the percentage of hits or misses for the model. Note that the model makes more correct predictions (hits, in green) when it is more certain of the prediction (range 0.87-0.96). As seen in Fig 5, this classifier yields a useful reliability profile with respect to its confidence score. This kind of characteristic means we can tune how many patients the model will indicate, as well as how sensitive or specific that indication can be. Such tuning can be tailored to any specific healthcare service, accounting for intensive care unit beds, available professionals and so on.

**Fig 5.**
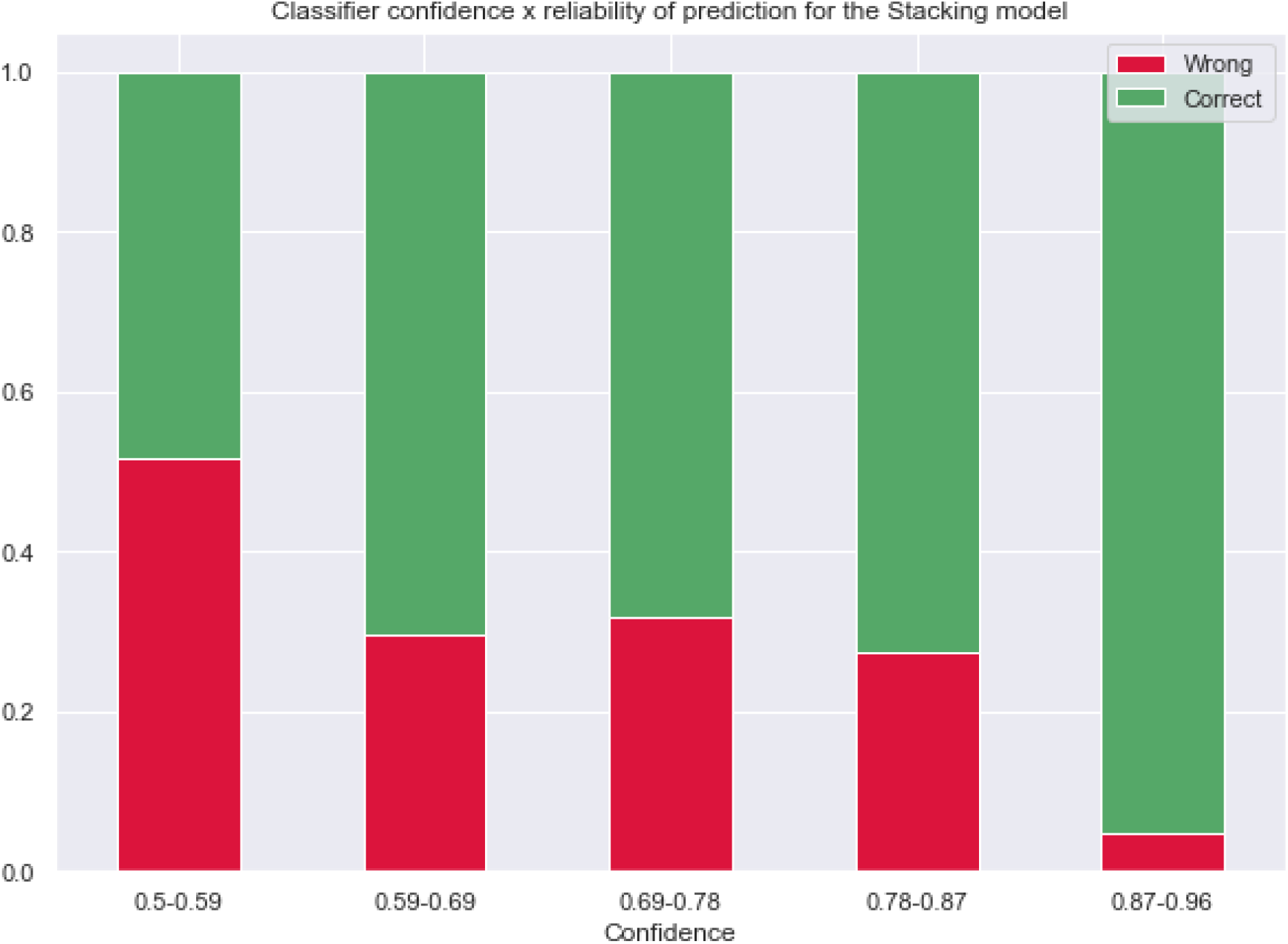
Error rates for each confidence threshold in the Stacking model.

In recent months, COVID-19 mortality prediction models were published ranging from simplified scores to machine learning. Based on S1 Table, there were few prediction studies that had extensive analysis utilizing AI techniques. In this study, AI techniques were compared to traditional statistical methods to develop a model to predict COVID-19 mortality, considering demographic, comorbidity, clinical presentation and laboratory analysis data. We observed that regarding the prediction of the class of interest (death), the best individual methods was a ML one (LightGBM) closely followed by a statistical model (GAM), both being better than neural network models, and both being surpassed by a meta-learning ensemble model -- Stacking -- which was the best overall solution considering all criteria for the posed prediction problem.

We would like to stress that, despite the fact that in medical research the AUROC is widely used as the sole measure of models’ discriminatory ability, our data reassured us that it is an insufficient metric for evaluating and comparing models. In contrast, F1 Score is a more robust metric, especially in larger, more complex and imbalanced datasets, which are common in health-related scenarios. Among the variables analyzed, age was the main mortality risk predictor, similar to other studies, while urea, C-reactive protein, lactate, respiratory rate, heart rate, NRL, neutrophils, sodium and pCO_2_ have been shown to significantly influence the disease outcome (according to Fig 3).

## Conclusion

In this study, modern AI techniques were compared to traditional statistical methods to develop a model to predict COVID-19 mortality with demographic, comorbidity, clinical presentation and laboratory analysis data. In our experiments, ML models excel in the task, with a meta-learning strategy based on Stacking surpassing the state-of-the-art LASSO regression method by more than 26% for predicting death. As a side effect of our study, we demonstrated that AUROC score was an insufficient metric for evaluating and comparing models. Even though some models, like Stacking and GAM have very similar AUROC scores, their capacity to discriminate relevant outcomes like death is quite different (0.53 F1 score for GAM and 0.56 for Stacking, which yields an 5.6% difference). Finally, we investigated issues related to the explainability and prediction reliability of the best ML models, concluding that they are potentially very useful for practical purposes in real settings.

## Data Availability

All data produced in the present study are available upon reasonable request to the authors.

## Acknowledgment

We would like to thank the hospitals which are part of this collaboration, for supporting this project: Hospital Bruno Born; Hospital Cristo Redentor; Hospital das Clínicas da Faculdade de Medicina de Botucatu; Hospital das Clínicas da UFMG; Hospital das Clínicas da Universidade Federal de Pernambuco; Hospital de Clínicas de Porto Alegre; Hospital Santo Antônio; Hospital Eduardo de Menezes; Hospital João XXIII; Hospital Julia Kubitschek; Hospital Mãe de Deus; Hospital Márcio Cunha; Hospital Mater Dei Betim-Contagem; Hospital Mater Dei Contorno; Hospital Mater Dei Santo Agostinho; Hospital Metropolitano Dr. Célio de Castro; Hospital Metropolitano Odilon Behrens; Hospital Moinhos de Vento; Hospital Nossa Senhora da Conceição; Hospital Regional Antônio Dias; Hospital Regional de Barbacena Dr. José Américo; Hospital Regional do Oeste; Hospital Risoleta Tolentino Neves; Hospital Santa Cruz; Hospital Santa Rosália; Hospital São João de Deus; Hospital São Lucas da PUCRS; Hospital Semper; Hospital SOS Cárdio; Hospital Tacchini; Hospital Unimed-BH; Hospital Universitário Canoas; Hospital Universitário Ciências Médicas; Hospital Universitário de Santa Maria.

We also thank all the clinical staff at those hospitals, who cared for the patients, and all undergraduate students who helped with data collection.

## Supporting information

**S1 Table. Main characteristics of the studies**. ARDS: acute respiratory distress syndrome; AST: aspartate transaminase; AUC: area under the curve; BMI: body mass index; BUN: blood urea nitrogen; CCEDRRN: canadian covid-19 emergency department rapid response network; CI: confidence interval; CKD: chronic kidney disease; COPD: chronic obstructive pulmonary disease; CPR: C-reactive protein; CT: computed tomography; DLN: deep learning networks; DM: diabetes mellitus; ED: emergency department; EH: emergency hospital; ER: emergency room; FiO_2_: fraction of inspired oxygen; GFR: glomerular filtration rate; GP: general practice; ICU: intensive care unit; IHD: ischemic heart disease; IL-6: interleukin 6; INR: international normalized ratio; LASSO: least absolute shrinkage and selection operator logistic regression; LDH: lactate dehydrogenase; MAP: mean arterial pressure; MICE: Multivariate Imputation by Chained Equations; NA: not applicable; NAAT: nucleic acid amplification test; NEWS2: national early warning score; NLR: neutrophil lymphocyte ratio; OP: outpatient; PLS: partial least squares; RDW: red blood cell distribution width; RF: Random Forest; RT-PCR: reverse transcription polymerase chain reaction; SF ratio: SpO_2_/FiO_2_ ratio; SVM: support-vector machine; Trop I: troponin I; XGBoost: eXtreme Gradient Boosting; WBC: white blood cell; WHO: World Health Organization.

**S2 Table. Potential predictors included for the development of the models**. BMI: body mass index; COPD: chronic obstructive pulmonary disease; HIV: human immunodeficiency viruses; pO_2_: partial pressure of oxygen; PCO_2_: partial pressure of carbon dioxide; SF ratio: SpO_2_/FiO_2_ ratio.

