## Supplementary material for "Effectiveness, Explainability and Reliability of Machine Meta-Learning Methods for Predicting Mortality in Patients with COVID-19: Results of the Brazilian COVID-19 Registry": Table 1

**Table S1. Main characteristics of the studies**

| **Study** | **Study design** | **Patient time span** | **Country of derivation** | **Country of validation** | **Sample size (n)** | **Development sample (n) (for mortality)** | **Validation sample (n) (for mortality)** | **Development population** | **Validation population** |
| --- | --- | --- | --- | --- | --- | --- | --- | --- | --- |
| Agarwal et al., | Retrospective observational study | Not clear | India | NA | 203 | 203 | NA | Hospitalized COVID-19 patients | NA |
| Alle et al., | Retrospective cohort | June 3^rd^ to October 23^rd^ 2020 | India | India | 544 | 429 | 115 | Patients with confirmed diagnosis of COVID-19 at MAX group of Hospitals, by 11 Sept 2020 | Patients with clear outcome after 11 Sept 2020 |
| Allenbach et al., | Prospective single-center cohort | March 16, 2020 to April 4, 2020 | France | France | 152 | 152 | 131 | Adult patients with confirmed COVID-19 from one tertiary care university hospital | Not described |
| Altschul et al., | Retrospective single-center cohort | March 1, 2020 to April 16, 2020 | United States of America | United States of America | 4711 | 2355 | 2356 | Patients with confirmed COVID-19 from an academic hospital | The same as the development population (spitted 50/50%, apparently by admission date) |
| Bello-Chavolla et al., | Registry data from an open source database from the Mexican Ministry of Health | First patient up to May 18, 2020 | Mexico | Mexico | 51633 | 41307 | 10326 | Patients with confirmed COVID-19 from the open source Mexican Ministry of Health database (inpatients and outpatients) | The same as the development population (split by random sampling stratified by mortality status) |
| Bertsimas et al., | Retrospective cohort | 02/01-05/15/2020 | Italy, Spain and United States of America | Greece, Spain and United States of America | 3,927 (external validation with 865 patients) | 2755 (training) | 307 | Adult patients admitted in the hospital, with confirmed SARS-CoV-2 infection by polymerase chain reaction testing of nasopharyngeal samples | The same (derivation population is randomly divided into training [85%] and testing [15%] set) |
| Booth et al., | Retrospective study | March 13, 2020 to June 5, 2020 (not clear) | United States of America | United States of America | 398 | 318 | 80 | Patients with positive RT-PCR assay results for SARS-CoV-2 | The same as the development population: training (80%) and testing (20%). |
| Chen et al., | Retrospective cohort | The first patient up to January 31, 2020 | China | China | 1590 | 1590 | NA | Patients with confirmed COVID-19 from 575 hospitals throughout China, excluding cases with incomplete medical records (20.8%) | NA |
| Chowdhury et al., | Retrospective study | 10 January to 18 February 2020 | China | China | 375 | 300 | 75 | Patients confirmed COVID-19 from a hospital in Wuhan | The same as the development population (80% data were used for training and validation while remaining 20% data were used for testing) |
| Churpek et al., | Observational study | March 4, 2020, and June 29, 2020 | United States of America | United States of America | 5075 | 3825 | 810 (independent validation) and 440 (validated the models from May 2020 to June 2020) | Adults with coronavirus disease 2019 admitted to 68 ICUs | 75% of the hospitals and tested in 25% of the hospitals |
| Dabbah et al., | Cohort study | March 16 2020 to December 12, 2020 | United Kingdom | United Kingdom | 7536 | 7536 | Not clear | Only participants with a positive RT-PCR COVID-19 test | Not clear |
| Das et al., | Retrospective cohort | January 20, 2020 to May 30, 2020 | South Korea | South Korea | 3524 | 3524 | NA | Data shared by Korea Centers for Disease Control and Prevention, from 17 provinces. Patients with confirmed COVID-19, with availability of demographic, exposure and diagnosis confirmation features along with the outcome | NA |
| El-Raheem et al. | Retrospective cohort study | November- December, 2020 | Sudan | NA | 105 | 105 | NA | COVID-19 patients attended in Hospital | NA |
| Faisal et al., | Registry data | March 11, 2020 to June 13, 2020 | United Kingdom | United Kingdom | 6444 | 3924 | 2520 | Consecutive adult non-elective or emergency medical admissions (COVID-19 and non-COVID-19 patients) from one hospital, who were discharged over a course of three months and had electronic NEWS2 recorded | Consecutive adult non-elective or emergency medical admissions (COVID-19 and non-COVID-19 patients) from another hospital, who were discharged over a course of three months and had electronic NEWS2 recorded |
| Fumagalli et al., | Retrospective cohort | February 22, 2020 to April 10, 2020 | Italy | Italy | 516 | 516 | NA | Consecutive adult patients with COVID-19 from 2 Italian tertiary hospitals | NA |
| Galloway et al., | Retrospective cohort | March 24, 2020 to April 17, 2020 | England | NA | 1157 | 1157 | NA | Patients with confirmed COVID-19 from 2 academic hospitals | NA |
| Gao et al., | Retrospective cohort | January 27, 2020 and March 21, 2020 | China | China | 2160 (917 external validation cohort) | 621 | 622 | COVID-19 patients with known outcomes (discharge or death) from hospitals Sino-French New City Campus of Tongji Hospital (SF), Optical Valley Campus of Tongji Hospital (OV) and The Central Hospital of Wuhan (CHWH) | We randomly partitioned 50 and 50% of participants from SF into training cohort (SFT cohort) and internal validation cohort (SFV cohort), respectively. |
| Garibaldi et al., | Retrospective cohort | March 4, 2020 to April 24, 2020, with follow-up through June 27, 2020 | United States of America | United States of America | 832 | 832 | NA | Consecutive confirmed COVID-19 patients from 5 hospitals (John Hopkins Medicine) | NA |
| Garrafa et al., | Retrospective cohort | March 2020 and December 2020 | Italy | Italy | 2782 | 1474 | 632 (tested on 676 second wave patients) | COVID-19 symptomatic patients, hospitalized during first wave | Same population (population divided into training [70%] and validation [30%] set) |
| Gavelli et al., | Retrospective single-center cohort | March 16, 2020 to April 22, 2020 | Italy | Italy | 480 | NA, it was developed by expert consensus | 480 | NA | Adult patients with confirmed COVID-19 patients admitted to one university hospital |
| Goméz et al., | Retrospective single-center cohort | February 24, 2020 to March 16, 2020 | Spain | NA | 163 | 163 | NA | Adult patients with suspected COVID-19 admitted to one university hospital | NA |
| Gue et al., | Retrospective single-center cohort | March 10, 2020 to May 30, 2020 | United Kingdom | NA | 316 | 316 | NA | Consecutive patients with confirmed COVID-19 from a general hospital, who had clinical symptoms at admission | NA |
| Hajifathalian et al., | Retrospective cohort | March 4, 2020 to April 9, 2020 | United States of America | United States of America | 929 | 664 | 265 | Adult patients with confirmed COVID-19 patients presenting to emergency department of 2 hospitals in Manhattan (did not exclude patients who were discharged within 24 hours) | Adult patients with confirmed COVID-19 patients presenting to emergency department of 9 hospitals in Massachusetts (did not exclude patients who were discharged within 24 hours) |
| Halalau et al., | Retrospective cohort | March 1, 2020 to April 1, 2020 | United States of America | United States of America | 2025 | Not clear | 1290 | Not clear | Confirmed SARS-CoV-2 patients who required hospital admission at 8 hospitals in Beamount, excluding patients who remained hospitalized beyond May 12, 2020 |
| Halasz et al., | Retrospective study | February to November 2020 | Italy | Italy | 852 | 596 | 256 | Patients (≥18 years) diagnosed with COVID-19 pneumonia (positive RT-PCR assay), admitted in hospital | Patients was randomly split in derivation (70%) and test (30%) cohorts |
| Hohl et al., | Observational study | March 1, 2020 and January 31, 2021 | Canada | Canada | 8761 | 6758 | 2054 | Patients with confirmed COVID-19 who presented to the ED of a participating site between March 1, 2020 and January 31, 2021. | The same as the development population. |
| Hou et al., | Retrospective study | February 7, 2020, and May 27, 2020 | United States of America | United States of America | 635 | 508 | 127 | Patients > 21 years old, tested positive for SARS-CoV-2 and admitted to the hospital | The same as the development population (80% training and 20% testing) |
| Hu et al., | Retrospective cohort | 28 January 2020 and 11 March 2020 | China | China | 247 | 183 | 64 | Patients with severe confirmed COVID-19 infection admitted to one hospital in Wuhan. Patients who had >10% missing values, stayed in the hospital <7 days, were afflicted by a severe disease before admission (e.g. cancer, aplastic anaemia or uraemia), were unconscious at admission or were directly admitted to the intensive care unit (ICU) were excluded. | The same as the development population, admitted at another hospital |
| Incerti et al., | Retrospective cohort study | February 20, 2020 and June 5, 2020 | United States of America | United States of America | 17086 | 13658 | 3428 | Patients were required to be older than 18 years old and have: (1) a U07.1 or U07.2 diagnosis, (2) a positive SARS-CoV-2 diagnostic test (e.g., either molecular or antigen tests) or (3) a B97.29 diagnosis with the absence of a negative SARS-CoV-2 molecular test within a 14-day window. | Patients were randomly assigned to either training (80%) or test (20%) sets. |
| Jamshidi et al., | Cohort study | February to September 2020 | Iran | NA | 23749 | Not clear | Not clear | Confirmed or suspected SARS-CoV-2 infections of people aged 18-100 years. | Four subsets were used as training data, and one subset was retained as a validation set for model testing |
| Kapoor et al., | Retrospective analysis | March 2020 to December 2020 | India | NA | 168 | 168 | NA | Lab-confirmed COVID-19 patients, with at least 18 years of age, from March 2020 till December 2020. | NA |
| Kazemi et al., | Retrospective cohort | February 25, 2020 to April 25, 2020 | Iran | NA | 91 | 91 | NA | Adult patients with confirmed COVID-19 who had undergone CT scan <8 days from the beginning of symptoms, excluding the ones with RT-PCR more than 7 days from CT. | NA |
| Kim et al., | Retrospective single-center cohort | February 19, 2020 to March 15, 2020 | Korea | NA | 38 | 38 | NA | Adult patients with confirmed COVID-19 admitted to a tertiary university hospital | NA |
| Knight et al., | Prospective cohort | May 21, 2020 to June, 29 2020 | England, Scotland and Wales | England, Scotland, and Wales | 57824 | 35463 | 22361 | Consecutive adult patients with COVID-19 from 260 hospitals, admitted up to May 20, 2020 | The same as the development population, admitted after May 20, 2020 |
| Ko et al., | Retrospective cohort | Development cohort: January 10, 2020 to February 24, 2020; Validation cohort: February to July 2020 | China | China | 467 | 361 | 106 | Patients with COVID-19 (not clear if laboratory-confirmed) from one hospital, excluding 14 patients without a blood test within 1 day after the hospital admission | Patients with COVID-19 (not clear if (laboratory-confirmed) from 3 hospitals |
| Le Lannou et al., | Prospective cohort | 30 January to 9 September 2020 | England | NA | 2499 | 2499 | NA | Patients (40 to 70 years) who are positive for COVID-19 from the Hospital Episode Statistics for England (HES) database | NA |
| Levy et al., | Retrospective and prospective cohort | March 1, 2020 to May 12, 2020 | United States of America | United States of America | 8391 | 6162 | 2229 | Adult patients with confirmed COVID-19 from 11 acute care hospitals in New York, from March 1, 2020 to April 23, 7 2020. Patients were excluded if they were still in the hospital at the study end point with a length of stay less than 7 days; if they were transferred to a hospital outside of the health system and their outcomes were unknown; or if they expired but were not marked as discharged in the EH | The same as the development cohort from another hospital in New York from March 1, 2020 to May, 7 2020, and all 12 hospitals from April 24, 2020 to May 6, 2020. |
| Li et al., | Retrospective study | 7 February 2020 and 4 May 2020 | United States of America | United States of America | 1108 | 997 | 111 | Patients diagnosed by positive tests of real-time polymerase chain reaction (RT-PCR) for SARS-CoV-2 | The same (data were split 90%  for training and 10% for testing) |
| Liang et al., | Retrospective cohort | November 21, 2019 to January 31, 2020 | China | China | 2300 | 1590 | 710 | Patients with COVID-19 from 575 hospitals in 31 provincial administrative regions | Data from hospitals not included in the development cohort |
| Lin et al., | Retrospective analysis | January 10, 2020, and February 24, 2020 | China | Korea | 464 | 361 | 103 | COVID-19 patients in Wuhan | COVID-19 patients in 3 Korean medical institutions |
| Lu et al., | Retrospective single-center cohort | January 21, 2020 to February 5, 2020 | China | NA | 577 | 577 | NA | Patients with confirmed or suspected COVID-19 from one hospital | NA |
| McRae et al., | Cohort study | March 1-October 30,2020 | Canada | Canada | 27665 | 21743 | 5922 | Patients presenting to 32 CCEDRRN sites that collected data on all patients tested for SARS-CoV-2. Consecutive eligible patients aged 18 and older who had a biological sample (swab, endotracheal aspirate, bronchoalveolar lavage) specimen collected for NAAT on their index emergency department visit or, if admitted, within 24h of emergency department arrival. For patients with multiple emergency department encounters involving COVID-19 testing, they only used the first encounter in this analysis. | Patients with same criterion (75% of eligible patients and outcome events to the derivation cohort and 25% to the validation cohort), but of geographically distinct. |
| Mei et al., | Retrospective cohort | January 21, 2020 to February 27, 2020 | China | China | 492 | 237 | Validation 1 = 120 and validation 2 = 135 | Adult patients with confirmed COVID-19, diagnosed with pneumonia by CT scan, from one hospital in Wuhan. Patients who died within the first 24 hours, with no clinical outcome available or who refused to participate were excluded. | The same as the development population, from other 3 hospitals |
| Momeni-Boroujeni et al., | Retrospective query | February 2020 until the end of March 2020 | United States of America | NA | 553 | 553 | NA | Patients admitted to SUNY Downstate Medical Center with COVID-19-related symptoms and confirmed Polymerase Chain Reaction (PCR)-positive | NA |
| Monterde et al., | Restrospective Study | June 15 to December 8 2020 | Spain | NA | 4607 | 4607 | NA | All COVID-19 hospitalizations reported in 46 eight public hospitals of Catalonia (North-East Spain) | NA |
| Naseem et al., | Restrospective Study | February 2020 and September 2020 | Pakistan | Pakistan | 1214 | 850 | 364 | Adult patients (≥18 years) admitted  to the Aga Khan University Hospital, Pakistan with a working diagnosis of COVID-19  infection | The same (final dataset was randomized and divided into training and testing sets with a 70/30% split respectively) |
| Nicholson et al., | Retrospective cohort | First patient to May 19, 2020 | United States of America | United States of America | 1042 | 578 | 464 | Consecutive adult patients with laboratory-confirmed COVID-19 patients from Mass General Brigham hospitals | Not clear |
| Núñez-Gil et al., | Retrospective cohort | February 8, 2020 to April 1, 2020 | Spain and Italy | NA | 908 | 908 | NA | Patients with confirmed COVID-19 from centers in Italy (n= 88) and Spain (n= 820) | NA |
| Sarkar et al., | Registry data | 13^th^ January, 2020 to 28^th^ February, 2020 | 22 countries in Asia, Australia, Europe and North America | NA | 115 | 115 | NA | Open source database of COVID-19 patients (inclusion criteria is not clear) | NA |
| Schlauch et al., | Retrospective cohort | March 2 to September 23, 2021 | United States of America | United States of America | 46971 | 23485 | 23485 | Patients hospitalized with COVID-19 | Patients were randomly split into training and testing sets with a 50:50 ratio |
| Shang et al., | Retrospective Cohort | January 1, 2020 to March 27, 2020 | China | China | 452 | 113 | 339 | Consecutive patients with confirmed COVID-19 from 2 hospitals in Wuhan, who had severe or critical illness | The same definition as the development population, but from a third hospital in Wuhan |
| Soto-Mota et al., | Retrospective Cohort | April 30, 2020 to May 20, 2020 | Mexico | NA | 400 | Score developed by consensus | 400 | NA | Consecutive patients with confirmed COVID-19 from 12 hospitals, with complete clinical information and outcome |
| Sottile et al., | Prospective and retrospective cohort | March 2020 and July 2020 | United States of America | United States of America | 27296 | 21837 | 5459 | All patients (with and without COVID-19) >14 years old hospitalized during the study period without a do not resuscitate order | COVID-19 positive cohorts |
| Sourij et al., | Prospective and retrospective cohort | April 15, 2020 to June 30, 2020 | Austria | NA | 238 | 238 | NA | Adult patients with confirmed COVID-19 and diabetes or pre-diabetes | NA |
| Stachel at al., | Retrospective cohort study | 3 March 2020–28 April 2020 | United States of America | United States of America | 3395 (external test set between 17–28 April: n=864) | 2054 | 477 | Patients admitted with COVID-19. There was used the time-holdout method and split hospital admissions into a training dataset (3 March–12 April: n=2054) | Patients confirmed with COVID: an internal validation dataset (13–16 April, n=477) |
| Turrini et al., | Retrospective cohort | Not clear | Italy | NA | 205 | 205 | NA | Patients aged between 17 and 100 years hospitalized for SARS-Cov-2 pneumonia | NA |
| Vaid et al., | Cohort study | Not clear | United States of America | Not clear | 4029 | Not clear | Not clear | COVID-19 positive patients | Not clear |
| Vaid et al., | Prospective and retrospective cohort | March 15 to May 22, 2020 | United States of America | United States of America | 4098 | 1514 | 383 (externally validated: 2201) >> 383 é validação temporal e 2201 geográfica? | Tested positive for COVID-19 | Patient clinical data from Mount Sinai Hospital (MSH) before the temporal split (May 1) were used to train and internally validate our XGBoost model in comparison with other baseline models. We then tested the series of XGBoost models on unimputed patient data on patients from four other external hospitals within the MSHS for external validation. |
| van Dam et al., | Retrospective analysis | 3 March and until 25 May 2020 | Netherlands | NA | 642 | 642 | Not clear | Adults admitted to the hospital with symptoms suggestive of COVID-19 and positive result of the PCR or (very) high suspicion of COVID-19 according to the chest CT scan | Not clear |
| Vela et al., | Retrospective study | March 1 and September 15, 2020 (development period), September 16 and December 27, 2020 (validation period) | Spain | Spain | 7718329 | 7.5 million | 218329 | Data from the entire population of Catalonia | Individuals with PCR-confirmed COVID-19, who were infected after developing the model |
| Wang et al., | Retrospective single-center cohort | January 28, 2020 to March 4, 2020 | China | China | 243 | 199 | 44 | Adult patients with confirmed COVID-19 from one university hospital | Data for enrolled patients were partitioned into two complementary subsets: the training set of patients from four wards was used to establish the predictive model, and the testing set of patients from another ward was used to validate the analysis. |
| Weng et al., | Retrospective cohort | January 1, 2020 to February 15, 2020 | China | China | 301 | 176 | 125 | Adult patients with laboratory-confirmed COVID-19 from 2 hospitals | The same as the development population (the criteria used to divide patients in training and testing sets was not clear) |
| Williams et al., | Retrospective cohort | Development cohort: any time prior to 2020; validation cohort: January 1^st^ 2020 to April 20, 2020 | United States of America, South Korea, Spain, Australia, Japan, Netherlands | South Korea, Spain, United States of America | 2126784 | 2082277 | 44507 | Healthcare database of 6 countries, in which adult patients with GP, EP or OP visit with influenza or flu-like symptoms, at least 365 days of prior observation, and no symptoms in the preceding 60 days | Adult patients with confirmed with COVID-19, presenting at an initial healthcare provider interaction in a GP, ER or OP visit, and who had no diagnosis of influenza or pneumonia and no flu-like symptoms in the preceding 60 days |
| Wollenstein-Betech et al., | Cohort study | As of May 1^st^, 2020 | Mexico | NA | ~91000 | ~91000 | No | Data from a publicly available repository, updated daily, containing information from approximately 91000 patients, as of May 1^st^, 2020 | No |
| Wu et al., | Prospective and retrospective cohort | 23 December 2019 to 13 February 2020 | China | China, Italy and Belgium | 725 | 239 | 60 | Patients with confirmed COVID-19 disease and presenting at hospital for admission | All patients included had the same inclusion and exclusion criteria, 80% for training (239 patients from 23 December 2019 to 28 January 2020) and 20% for internal validation (60 patients from 29 January to 13 February 2020). The external validation was collected between 20 February 2020 and 31 March 2020 from centres in China, Italy and Belgium under the same inclusion and exclusion criteria. |
| Xie et al., | Retrospective cohort | January and February 2020 | China | China | 444 | 299 | 145 | Patients with confirmed COVID-19 from one hospital in Wuhan who had been discharged or died | Patients with confirmed COVID-19 from another hospital in Wuhan, excluding 6 patients who died quickly |
| Yadaw et al., | Retrospective and prospective cohort | March 9, 2020 to April 7, 2020 | United States of America | United States of America | 5051 | 3841 | 961 | Inpatients and outpatients (including those attended by telehealth) with confirmed COVID-19 from the Mont Sinai Health System (8 hospitals and over 400 ambulatory practices) until April 6, 2020 | The same as the development population (randomly split 80/20%) and patients admitted to Mont Sinai Hospitals who were included in the database (with the outcome) on April 7, 2020 |
| Yan et al., | Retrospective cohort | Development cohort: January 10, 2020 to February 18, 2020; Validation cohort: February 19-24, 2020 | China | China | 485 | 375 | 110 | Adult patients with COVID-19 (not clear if patients had laboratory-confirmed disease), from one hospital, excluding patients with >20% missing values and breast-feeding women | The same as the development population, admitted after February 18, 2020 |
| Yoo et al., | Retrospective cohort | March 1, 2020 to April 28, 2020 | United States of America | United States of America | 4840 | 1613 | 1614 | Adult patients with confirmed COVID-19 from 5 hospitals, up to 99 years-old. The sample was randomly split in 3 datasets, the second one was used for development | The same as the development population: randomly split in 3 datasets, the third one was used for validation |
| Yu et al., | Retrospective multicenter study | 2/1/2020 and 5/4/2020 | United States of America | United States of America | 3491 | 2793 | 698 | All patients with COVID-19 patients, who presented to the ER at Beaumont Health, consisting of eight hospitals | The patient cohort was randomly split into training (80%) and testing (20%) groups to train and test CatBoost model. |
| Zhang et al., | Retrospective cohort | January 12, 2020 to February 9, 2020 | China | China | 828 | 516 | 312 | Adult patients with confirmed COVID-19 from one hospital | Adult patients with confirmed COVID-19 from the same hospital in a different time span (February 8-9, 2020) and from another hospital |
| Zhang et al., | Retrospective cohort | Not reported | China | United Kingdom | 1001 | 775 | 226 | Adult patients with confirmed COVID-19 from one hospital | Adult patients with confirmed COVID-19 from another hospital |
| Zhao et al., | Retrospective study | March 9, 2020 to April 20, 2020. | United States of America | United States of America | 641 | 454 | 187 | Hospitalized patients with laboratory-confirmed COVID-19, with age ≥ 18 years old | The same (70% for  training and 30% for testing) |
| Zhou et al., | Retrospective, multicentre cohort study | Dec 29, 2019 and Jan 31, 2020 | China | NA | 191 | 191 | No | All adult inpatients (≥18 years old) with laboratory confirmed COVID-19 from Jinyintan Hospital and Wuhan Pulmonary Hospital (Wuhan, China) who had been discharged or had died by Jan 31, 2020. | No |
| Zhou et al., | Retrospective single-center cohort | January 12, 2020 to February 26, 2020 | China | NA | 118 | 118 | NA | Elderly patients (>60 years) with "clinically diagnosed" COVID-19 (RT-PCR or chest CT) from one university hospital | NA |
| Zhu et al., | Retrospective Study | January 29, 2020 to March 21, 2020 | China | China | 181 | 154 | 27 | Patients with confirmed COVID‐19 infection | The same as the development population (data were split into 85% training and 15% testing) |

**Table 5. Continued**

| **Author** | **Model outcome** | **Outcome time** | **Original modelling approach** | **Imputation** | **Use of AI techniques** | **Number of variables were tested in the development cohort** | **Variables included in the final model (for mortality)** |
| --- | --- | --- | --- | --- | --- | --- | --- |
| Agarwal et al., | Severity and mortality | In-hospital | Multivariate logistic regression | No | No | 10 | Absolute neutrophil count, neutrophil to lymphocyte ratio means, Early lymphocyte count |
| Alle et al., | Risk stratification and mortality | In-hospital | Machine Learning and logistic regression model | Yes, KNN imputation | Yes, XGboost, Random Forest and SVM | 357 | D-Dimer, 328 Ferritin, Lymphocyte (%), Neutrophil to Lymphocyte ratio (NLR), WBC, Trop I, INR, IL-6 and LDH |
| Allenbach et al., | Composite of ICU admission or death | 14 days | Multivariate logistic regression | No | No | 42 | Age, WHO clinical scale, CRP and lymphocytes count obtained on admission |
| Altschul et al., | Mortality | In-hospital | Multivariate logistic regression | No | No | Not clear | Age, sex, SpO_2_, MAP, INR, creatinine, BUN, IL-6, CRP and procalcitonin obtained on admission |
| Bello-Chavolla et al., | Mortality | 30 days | Cox proportional risk regression analysis | No | No | 12 | Age, diabetes, obesity, CKD, COPD, hypertension, immunosuppression and COVID-19 pneumonia |
| Bertsimas et al., | Mortality | In-hospital | Machine Learning | Yes, k-nearest neighbors imputation | Yes, XGBoost | 22 | Increased age, decreased oxygen saturation (≤ 93%), elevated levels of C-reactive protein (≥ 130 mg/L), blood urea nitrogen (≥ 18 mg/dL), and blood creatinine (≥ 1.2 mg/dL) were identified as primary risk factors |
| Booth et al., | Mortality | In-hospital and post-discharge | Machine learning techniques | Yes, scikit-learn’s Iterative Imputer method | Yes, support vector machine | 26 | C-reactive protein, blood urea nitrogen, serum calcium, serum albumin, and lactic acid. |
| Chen et al., | Mortality | 14, 21 and 28 days | Multivariate Cox regression analysis | No | No | 37 | Age, coronary heart disease, cerebrovascular disease, dyspnea, procalcitonin, aspartate aminotransferase, total bilirubin upon admission |
| Chowdhury et al., | Mortality | In-hospital | Machine learning techniques | Yes, MICE and (− 1) | Yes, Multi-tree XGBoost model | 76 | Lactate dehydrogenase, neutrophils (%), lymphocyte (%), high-sensitivity C-reactive protein, and age (LNLCA). |
| Churpek et al., | Mortality | In-hospital | Machine Learning | Yes, Bagged trees using the caret package in R (The R Foundation for Statistical Computing, Vienna, Austria) | Yes, Elastic Net Logistic Regression, eXtreme Gradient Boosting, Random Forests, Neural Networks, Support Vector Machines and K-Nearest Neighbors | 64 | Age, vital signs and respiratory support on ICU admission, Fio2 among patients requiring invasive mechanical  ventilation, laboratory values, and organ support. |
| Dabbah et al., | Mortality | Especially in hospital-at-home settings | Random Forest (RF) and Cox models | No | Yes, Random Forest (RF) | 100 top-ranked features out of ~12000 feature | 3 vital signs; 11 symptoms; 32 pre-existing clinical conditions; 5 medications and treatments; and 13 patient characteristics (Table 1). The top risk factors  were age, acute kidney failure (<1 month), and waist circumference (Figure 3). |
| Das et al., | Mortality | In-hospital | Logistic regression and machine learning techniques | No | Yes. SVM, K nearest neighbor, RFM and gradient boosting | 4 | Age, sex, province (in South Korea) and exposure (nursing home, hospital, religious gathering, call center, community center, shelter and apartment, gym facility, overseas inflow, contact with patients and others) |
| El-Raheem et al., | Mortality | In-hospital | Logistic regression model | No | No | 21 | Age, gender, admission, IHD, fever, cough, complications, duration of hospitalization and enoxaparin dose |
| Faisal et al., | Mortality | In-hospital | Multivariable logistic regression | No | No | Not clear | CARMc19_N: 10 [age, sex, COVID-19 (yes/no), NEWS2 score and subcomponents] and CARMc19_NB: 18. All variables from CARMc19_N + 7 blood test results + AKI score |
| Fumagalli et al., | Mortality | In-hospital | Cox regression analysis | No | No | 20 | Age, number of comorbidities (CV disease, hypertension, DM, depression, dementia and cancer), respiratory rate, PaO2/FiO2, serum creatinine and platelet count obtained on admission |
| Galloway et al., | Composite of transfer to ICU or death | In-hospital | LASSO logistic regression | No | No | 19 | Age, sex, ethnicity, DM, hypertension, chronic lung disease, SpO2, radiographic severity score, neutrophil count, respiratory rate, CRP, albumin, creatinine obtained on admission |
| Gao et al., | Mortality | In-hospital | Machine learning techniques | Yes, R-package missForest and random forest | Yes, Support Vector Machine, Gradient Boosted Decision Tree, and Neural Network | 53 | Age, blood urea nitrogen [BUN], respiratory rate [RR], D— dimer, age, platelet count [PLT], albumin [ALB], SpO2 and lymphocyte |
| Garibaldi et al., | In-hospital mortality and a composite of disease severity (WHO scale) or in-hospital mortality | In-hospital | Cox regression analysis | Yes. Imputed missing values by chained equations (MICE) with predictive mean matching | Yes. NLP was used to identify presenting symptoms | 24 | Age, nursing home residence, sex, BMI, Charlson Comorbidity Index, SaO2/FiO2 ratio obtained on admission |
| Garrafa et al., | Mortality | In-hospital | Machine Learning and Logistic Regression | Yes, “on-the-fly-imputation” algorithm | Yes, Random Forests and Gradient Boosting Machine (GBM) | 22 | Age (the most powerful predictor), blood analytes (the strongest predictors were lactate dehydrogenase, D-dimer, Neutrophil/Lymphocyte ratio, C-reactive protein, Lymphocyte %, Ferritin std and Monocyte %), and Brescia chest X-ray score |
| Gavelli et al., | In-hospital mortality and in-hospital clinical stability | In-hospital | Multivariable logistic regression and Cox Regression Hazard models | No | No | NA | Presence of comorbidity (any disease on active therapy), SpO2 and respiratory rate after a trial of 15 minutes with oxygen at a FiO2 0.5 |
| Goméz et al., | Mortality | 30 days | Multivariable logistic regression | No | No | 20 | Age, creatine, glucose and white blood cells obtained on admission |
| Gue et al., | Mortality | 30 days | Multivariable logistic regression | No | No | 15 | Age, sex, hypertension, coronary artery disease, heart failure, atrial fibrilation, oral anticoagulants, modified sepsis-induced coagulopathy (mSIC) score (INR, platelet count, qSOFA score) |
| Hajifathalian et al., | Mortality | 7 days and 14 days | Multivariable logistic regression | Yes. Imputation by chained equations | No | 38 | Age, mean arterial pressure, serum creatinine and severity of hypoxia at hospital presentation |
| Halalau et al., | Hospital admission and in-hospital mortality | In-hospital | Multivariate logistic regression | No | No | Not clear | Age, male sex, congestive heart failure, end-stage renal disease, chronic pulmonary disease, DM, hypertension, obesity, nursing home residence, immunocompromised status, congenital heart disease, coronary artery disease, end-stage liver disease and pregnancy |
| Halasz et al., | Mortality | In-hospital | Machine learning algorithm | No | Yes, Naive Bayes and bootstrapping | 62 | Age; mean corpuscular haemoglobin concentration; PaO2/FiO2 ratio; temperature; previous stroke; gender |
| Hohl et al., | all-cause Emergency Department and in-hospital mortality. | In-hospital | Logistic Regression | Yes, 5 multiple imputations for predictors if missing categorical data could not reasonably be assumed to be absent. Authors validated the model in a cohort of geographically distinct sites that were not part of derivation, and used a single imputation for the few missing respiratory rates (4% were missing). | No | 41 | Age, sex, arrival from, Arrival Mode, Chest Pain, Moderate/Severe Liver disease, Arrival Respiratory Rate ǂ, Mode and Level of Oxygen in ED |
| Hou et al., | Intensive care unit admission and mortality | In-hospital | Machine learning | Yes, Multivariate Imputation by Chained Equations. No imputation was performed for a predictor with missing >15%. Brain natriuretic peptide and troponin had more than 30% missing and were excluded from the analyses. | Yes, Random forest, Xgboost, kernel support vector machine and neural network | 29 | Age, procalcitonin, C-reactive protein, lactate dehydrogenase, D-dimer and lymphocyte |
| Hu et al., | Mortality | In-hospital | LASSO logistic regression | Yes, using bagging tree. Variables with >30% missing values were excluded | Yes. Logistic regression, PLS regression, EN model, random forest and bagged flexible discriminant analysis (FDA). | 51 | Age, CRP, D-dimer, lymphocyte count at admission |
| Incerti et al., | Mortality | In-patient | Multivariable logistic regression | Yes, Multivariate imputation by chained equation | No | 55 | Age, oxygen saturation, temperature, respiratory rate, lactate dehydrogenase, and white blood cell count |
| Jamshidi et al., | Symptom prediction and mortality | In-hospital | Machine learning | No | Yes,  Random Forest, Artificial Neural Network, K-Nearest Neighbors, Linear Discriminant Analysis and Naive Bayes. | 15 | Respiratory distress, consciousness disorders, chest pain, paresis or paralysis, cough, fever or chill, gastrointestinal symptoms, sore throat, headache, vertigo, loss of smell or taste, and muscular pain or fatigue |
| Kapoor et al., | COVID-19 disease severity, in-hospital mortality, and pulmonary embolism risk | In-hospital | Logistic regression | NA | No | 127 | Age, Duration of Hospital stay (days), Temperature, Respiratory Rate, Oxygen Saturation, Systolic Blood Pressure, Diastolic Blood Pressure, heart rate, Total Leucocyte Count, Hemoglobin, D-Dimer, Fibrinogen, Platelet count, DIC score, DIC score, Prothrombin time; International Normalized Ratio, CT severity index |
| Kazemi et al., | Mortality | In-hospital | Multivariate logistic regression | No | No | Not available | Age, sex, comorbidity (cardiovascular and pulmonary), diffused distribution of CT abnormality, total CT-score and dyspnea at admission |
| Kim et al., | Mortality | In-hospital | Statistic | No | No | 3 | Myocardial damage marker (creatine kinase-MB [CK-MB] or troponin-I > the 99th percentile upper reference limit) + Heart failure marker (NT-proBNP ≥ 125 pg/mL) + Electrical abnormality marker (first detected or newly developed supraventricular tachycardia, ventricular tachycardia, ventricular fibrillation, atrial fibrillation, bundle branch block, ST-segment elevation/depression, T-wave flattening/inversion, and QT interval prolongation on ECG) |
| Knight et al., | Mortality | In-hospital | LASSO logistic regression | Yes. Multiple imputation with chained equations | Yes. XGBoost | 41 | Age, sex, number of comorbidities (chronic cardiac disease, chronic respiratory disease excluding asthma, chronic renal disease defined as estimated glomerular filtration rate ≤30, mild to severe liver disease, dementia, chronic neurological conditions, connective tissue disease, DM, HIV or AIDS, and malignancy), respiratory rate, SpO2, level of consciousness, urea and CPR obtained on admission |
| Ko et al., | Mortality | In-hospital | Machine learning techniques | Yes, imputed with mean values for development and training datasets | Yes, deep neural network and random forest models | 73 | Lymphocytes, neutrophils, albumin, LDH, neutrophil count (?), CRP, prothrombin activity, calcium, urea, estimated GFR, monocytes, globulin, eosinophils, glucose, RDW, bicarbonate, RDW standard deviation, platelet count, mean platelet volume, platelet large-cell ratio, prothrombin time, total protein, platelet distribution width, aspartate aminotransferase, thrombocytocrit, eosinophil count, alkaline phosphatase, INR |
| Le Lannou et al., | Mortality | In-hospital | Machine Learning | No | Yes, Random Forest | 26 | Age, diabetes, renal disease, hypertension, respiratory diseases, sex, predisposition to infection and cardiovascular diseases |
| Levy et al., | Mortality | 7 days | LASSO logistic regression | Yes, imputation of means. Variables with >50% missing values were excluded. | No | 42 | Age, length of stay, SpO2, neutrophil, RDW, sodium urea (on admission and every 2 days) |
| Li et al., | ICU admission and mortality | In-hospital | Machine learning | Yes, MICE | Yes, deep neural network mode | 55 | Age, lactate dehydrogenase, procalcitonin, cardiac troponin, C-reactive protein and oxygen saturation |
| Liang et al., | Composite of ICU admission, need of invasive mechanical ventilation or death | In-hospital | LASSO logistic regression | Yes (if <20%). Predictive mean matching to impute numeric features, logistic regression to impute binary variables, and Bayesian polytomous regression to impute factor features | No | 72 | Chest radiographic abnormality, age, hemoptysis, dyspnea, unconsciousness, number of comorbidities (COPD, hypertension, DM, coronary heart disease, chronic kidney disease, cancer, cerebrovascular disease, hepatitis B, immunodeficiency), cancer history, neutrophil-to-lymphocyte ratio, lactate dehydrogenase and direct bilirubin obtained on admission |
| Lin et al., | Mortality | In-hospital | Machine learning | No | Yes, artificial neural network (ANN) and convolutional neural network (CNN) | 30 | More important: lymphocyte, lactate dehydrogenase and high sensitivity C-reactive protein |
| Lu et al., | Mortality | 12 days | Cox regression analysis | No | No | Not clear | Age, CPR |
| McRae et al., | Probability of SARS-CoV-2 infection without the need for laboratory testing | In-hospital | Logistic regression model with continuous predictors fit with restricted cubic splines with three knots. The strengths of associations between predictors and outcome were assessed using an analysis of variance (ANOVA). Internal bootstrap validation with 1,000 bootstrap samples was conducted to provide an optimism-corrected C-statistic. We then developed the points-based CCEDRRN COVID-19 Infection Score (CCIS) using a nomogram. Discrimination of the score was evaluated using the C-statistic. | Five multiple imputations were used for continuous variables with missing data. | No | 45 | 7-day average incident COVID-19 cases, Institutional exposure (e·g· Long-term care, prison) or Travel from country with known cases within 14 days, Healthcare worker/Microbiology lab, Household/caregiver contact, Temperature, Supplemental oxygen delivered in the Emergency Department, Cough, Dysgeusia/Anosmia, Muscle aches (Myalgia), Current tobacco user |
| Mei et al., | Mortality | In-hospital | LASSO logistic regression | No | No | 43 | Age, NLR, admission body temperature, AST and, total protein |
| Momeni-Boroujeni et al., | Mortality | In-hospital | Markov model and Regression Logistic | No | No | 50 | Age >80 years, history of coronary artery disease and chronic obstructive pulmonary disease increased mortality risk. The lab values upon admission most associated with mortality included neutrophil percentage, red blood cells (RBC), red cell distribution width (RDW), protein levels, platelets count, albumin levels and mean corpuscular hemoglobin concentration (MCHC). |
| Monterde et al., | Critical illness in hospitalized COVID-19 patients was a composite that included the need for invasive mechanical ventilation, transfer to the intensive care unit (ICU), or in-hospital death | In-hospital | Logistic regression models | No | No | Not clear | Age, age group, sex, Charlson index, Elixhauser index, Queralt DxS index and Risk groups. |
| Naseem et al., | Mortality | In-hospital | Machine learning techniques | No | Yes, Deep-Neo-V model, Random Trees (CART), K-Nearest Neighbor (KNN), Support Vector Classifier - Radial Basis Function (SVC - RBF), Ada-Boost-Classifier (ABC) and Quadratic Discriminant Analysis (QDA) and a deep neural network (DNN) | 80 | Age, chronic obstructive lung disease [COPD], chronic kidney disease [CKD], ischemic heart disease [IHD]), pneumothorax (radiological or clinical diagnosis), acute respiratory syndrome [ARDS], septic shock, shortness of breath, ICU admission, AB+ Blood group and recurrent admission to the ICU, abnormalities like creatinine, blood urea nitrogen), INR and PT, invasive ventilation and non-invasive ventilation, having fever and the use of systemic steroids. |
| Nicholson et al., | Need of mechanical ventilation and in-hospital mortality | In-hospital | Multivariate logistic regression | No | No | 49 | Age, sex, diabetes mellitus, chronic statin use, albumin, C-reactive protein, neutrophil-lymphocyte ratio, mean corpuscular volume, platelet count, and procalcitonin obtained on admission |
| Núñez-Gil et al., | Mortality | In-hospital | Multivariate logistic regression | No | No | Not clear | Age, hypertension, obesity, renal insufficiency, any immunosuppressive condition, SpO2, CRP obtained on admission |
| Sarkar et al., | Mortality | In-hospital | Machine learning techniques | No | Yes, Random Forest | 6 | Age, sex, from Wuhan, visit to Wuhan, days from symptom onset to hospitalization |
| Schlauch et al., | Mortality | In-hospital | Logistic regression model | Yes, MICE | No | 49 | Not clear: age, WHO PS, male sex, comorbidities, and markers of inflammation and organ dysfunction, but also shows clear dose responses for critical laboratory measures |
| Shang et al., | Mortality | In-hospital | LASSO logistic regression | Yes, multiple imputation methods for variables with <10% missing values | No | 52 | Age, coronary heart disease, % of lymphocytes, procalcitonin and D-dimer |
| Soto-Mota et al., | Mortality | In-hospital | Consensus | No | No | NA | Age, hypertension, white blood cell count, lymphocyte count, myocardial necrosis marker, creatinine, SpO2 (not clear in which moment) |
| Sottile et al., | Mortality | In-hospital | Logistic regression models, estimating the stacked model, and evaluating the stacked mode | Yes | No | Not clear | Male, Hispanic, receive ICU-level care, be intubated, have a longer duration of mechanical ventilation, a longer hospital length of stay, and not survive. Patients with COVID-19 had higher SOFA and CURB-65 scores and LDH, ferritin, and D-dimer levels. Mean troponin levels were lower in patients with COVID-19 |
| Sourij et al., | Mortality | In-hospital | Multivariate logistic regression | No | No | Not clear | Age, arterial occlusive disease, CRP, estimated GFR and aspartate AST levels obtained on admission |
| Stachel at al., | Mortality | In-hospital | Machine learning | Yes, For LR, SVM and NN, missing values were imputed on  datasets using median values from observations found in  the training set in order to avoid dropping incomplete  cases and improve model training. For binary or categorical variables, the median was rounded to the nearest  integer. For DT and GB, missing values were treated as  separate values and used in the calculation of the worth  of a splitting rule. This consequently produces a splitting  rule that assigns the missing values to the branch that  maximises the worth of the split. This can be a desirable  option as existence of a missing values such as lab test can  be predictive of mortality. | Yes, decision tree, gradient boosting decision trees, support vector machine and neural network | 83 | Pulse, oximetry, respirations, systolic blood pressure, diastolic blood pressure, blood urea nitrogen, white blood cell, age, length of stay, lymphocyte, temperature, calcium, glucose, prothrombin time, phosphorous, neutrophils per cent, anion gap, troponin value and neutrophil (for complete description to see Figure 4) |
| Turrini et al., | Mortality | In-hospital | COX univariate and multivariate logistic regression | No | No | 14 | Age, number of relevant comorbidities, P/F ratio less than 200 at presentation, high levels of LDH, and elevated C-reactive protein (CRP) values |
| Vaid et al., | Mortality | In-hospital | Multilayer perceptron (MLP) and logistic regression with L1-regularization, or least absolute shrinkage and selection operator (LASSO). | No | Yes, Multilayer Perceptron | 31 | Not clear |
| Vaid et al., | Mortality and critical events at time windows of 3, 5, 7, and 10 days from admission | In-hospital | Machine learning | Yes, features with >30% missingness were dropped, and k-nearest neighbors (kNN, k=5) was used to impute missing data in the remaining feature space. To further assess the impact of imputation on performance, an XGBoost model was also created and trained on the imputed data set. Imputation for the training set (ie, MSH only) and external validation set (ie, OH) were performed using only the first collected value from the respective sites to prevent information leakage that could compromise assessment of generalizability. We assessed the calibration of the results of each model to ensure that the model predictions could be interpreted as real-world risk scores. Calibration was performed using both the sigmoid and isotonic methods of the CalibratedClassifierCV class in scikit-learn and evaluated using the Brier score metric. | Yes, Extreme Gradient Boosting | 52 | Age, Anion Gap, C-Reactive Protein, Lactate Dehydrogenase (LDH), Oxygen Saturation. Blood Urea Nitrogen, Ferriti, Red Cell Distribution Width (RDW), Diastolic Blood Pressure and Lactate |
| Van Dam et al., | Mortality and admission to intensive care unit | In-hospital | Logistic regression | Yes, multiple imputation | No | ±16 | P(30-day mortality)=1/(1+exp (-(−3.908+0.050*Age +1.115*≥2 Abnormal Vital Signs (yes=1, no=0)–0.112*Albumin (in g/L)+0.284*(BUN (in mmol/L)/5)+0.120*(LDH(in U/L)/100)+0.875*Bilirubin>20 µmol/L (yes=1,  no=0))) |
| Vela et al., | Hospital admission, transfer to intensive care unit (ICU), and death. | In-hospital | Generalized linear models (Poisson regression) | No | No | NA | Age and underlying conditions such as diabetes, arterial hypertension, and cardiovascular diseases, however, the comorbidity burden was a stronger predictor of deaths |
| Wang et al., | Mortality | 28 days | Multivariable logistic regression | No | No | 41 | Age, ferritin and D-dimer obtained on admission |
| Weng et al., | Mortality | In-hospital | LASSO logistic regression | Yes, for variables with <10% missing values (>10% were excluded from model development). RF. | No | 24 | Age, neutrophil-to-lymphocyte ratio, D-dimer and C-reactive protein obtained on admission |
| Williams et al., | Hospitalization with pneumonia, hospitalization with pneumonia requiring intensive services or death and death in the 30 days after index date | In-hospital and 30 days after index rate | LASSO logistic regression | No | No | 31917 | Age, sex, history of cancer, COPD, diabetes, heart disease, hypertension, hyperlipidemia and kidney disease |
| Wollenstein-Betech et al., | (1) hospitalization, (2) mortality, (3) need for ICU, and (4) need for a ventilator | In-hospital | Machine learning and logistic regression | No | Yes, sparse Support Vector Machines (SVM), sparse Logistic Regression (LR), Random Forests (RF) and gradient boosted decision trees (XGBoost). | Not clear | Age, SARS-CoV-2 test status, immunosuppression and pregnancy |
| Wu et al., | Severity risk assessment and triage for COVID-19 patients at hospital admission | In-hospital | Machine-learning model, nomogram and online calculator | No | Yes, boruta algorithm that combines feature rank based on the random forest classification algorithm and selection frequency based on multiple iterations of the feature selection procedure | 71 | Elderly patients, of male sex, non-hospital staff, suffering from hypertension, diabetes, cardiopathy disease, COPD, cerebrovascular disease, renal disease, hepatitis B virus infection, lower body temperature and chest tightness |
| Xie et al., | Mortality | In-hospital | Multivariate logistic regression | No | No | 28 | Age, lymphocyte count, lactate dehydrogenase and SpO2 obtained on admission |
| Yadaw et al., | Mortality | In-hospital | Artificial intelligence techniques | Yes, using means | Yes. Recursive feature elimination method for feature selection, and logistic regression, SVM, RF model, and XGBoost algorithms for prediction | 17 | 17F: age, sex, ethnicity, encounter type, temperature, diastolic blood pressure, oxygen saturation at presentation, minimum oxygen saturation, smoking, asthma, COPD, obesity, DM, HIV, cancer; 3F: age, minimum oxygen saturation, and type of patient encounter, obtained the day of admission |
| Yan et al., | Mortality | In-hospital | Machine learning techniques | No | Yes, XGBoost machine learning algorithm | 75 | LDH, lymphocytes and CRP obtained at hospital admission |
| Yoo et al., | Mortality | In-hospital | Gray`s K-sample tests, DeLong's test | No | No | 48 | Glasgow coma scale, oxygen support level, BUN, age, lymphocyte percentage and troponin |
| Yu et al., | Risk of invasive mechanical ventilation and mortality | In-hospital | Machine learning techniques | No | Yes, catBoost algorithms | 34 | Age, sex, race, BMI, smoking history, alcohol history, history of DM, history of lung disease, history of heart disease and history of kidney disease |
| Zhang et al., | Mortality | 14 days and 28 days | Cox regression analyses | Yes. Multiple imputations (method not reported) | No | 30 | Age, LDH, NLR and direct bilirubin obtained on admission |
| Zhang et al., | Death and poor outcome (developing ARDS, receiving intubation ou ECMO treatment, ICU admission or death) | In-hospital | LASSO logistic regression | No | No | 19 | DCS (demographic, comorbidities and symptoms): age, sex, chronic lung disease, DM, hypertension, immunosuppression, cancer, CKD, heart disease, cough, dyspnea, diarrhea; DCSL (demographic, comorbidities, symptoms and laboratory tests): age, sex, chronic lung disease, DM, cancer, cough, dyspnea, CRP, creatinine, platelets, neutrophils and lymphocytes counts; DL (demographic and laboratory tests): age, sex, CRP, creatinine, platelets, neutrophils and lymphocytes counts (around admission) |
| Zhao et al., | Intensive care unit (ICU) admission and mortality | In-hospital | Logistic regression | No | No | 49 | Heart failure, procalcitonin, lactate dehydrogenase, chronic obstructive pulmonary disease, pulse oxygen saturation, heart rate, and age |
| Zhou et al., | Mortality | In-hospital | Univariable and multivariable logistic regression methods | No | No | 43 | Older age, higher Sequential Organ Failure Assessment (SOFA) score and D-dimer greater than 1 µg/mL on admission |
| Zhou et al., | Mortality | In-hospital | Multivariable logistic regression | No | No | 37 | Lactate dehydrogenase, albumin, BUN, NLR and D-dimer obtained on admission |
| Zhu et al., | Mortality | In-hospital | Machine learning | Yes, Multivariate Imputation by Chained Equations in R, moreover the brain natriuretic peptide was removed from the dataset, for being missing in >15% of the patients. Collinearity analysis in feature selection was used to remove correlated variables. | Yes, deep neural network | 78 | D-dimer, O_2_ Index, neutrophil:lymphocyte ratio, C-reactive protein, and lactate dehydrogenase. |

**Table 5. Continued**

| **Study** | **How many patients died in the development dataset?** | **External validation** | **AUC in derivation cohort** | **AUC in validation cohort** | **F1 score** | **TRIPOD** |
| --- | --- | --- | --- | --- | --- | --- |
| Agarwal et al., | 51 | No | Not clear | NA | Yes | No |
| Alle et al., | 61 | No | (0.927±0.01) | Not clear | Yes | No |
| Allenbach et al., | 32 | No | 0.786 for the composite outcome and 0.803 for death (after correction for over-optimism; IC95% not reported) | 0.787 for the composite outcome and 0.827 for death (after correction for over-optimism; IC95% not reported) | No | Yes |
| Altschul et al., | 621 | Yes | 0.824 (0.814 to 0.851) | 0.798 (0.789 to 0.818) | No | No |
| Bello-Chavolla et al., | 4276 | Yes | 0.823 (95% CI not reported) | 0.830 (95% CI not reported) | No | No |
| Bertsimas et al., | 1054 | Yes | 0.90 (95% CI, 0.87–0.94) | 0.92 (95% CI, 0.88–0.95) on Seville patients, 0.87 (95% CI, 0.84–0.91) on Hellenic COVID-19 Study Group patients, and 0.81 (95% CI, 0.76–0.85) on Hartford Hospital patients | No | No |
| Booth et al., | 43 | No | 0.93 | Not available | No | No |
| Chen et al., | 50 | No | 0.91 (95% CI, 0.85-0.97) | NA | No | No |
| Chowdhury et al., | 174 | Yes | 0.961 | 0.991 | No | No |
| Churpek et al., | 1846 | Yes | Not clear | XGBoost mode: 0.81 (95% CI, 0.78–0.85) | No | No |
| Dabbah et al., | 496 | No | 0.92 | Not clear | Yes | Yes |
| Das et al., | 74 | No | 0.830 (95% CI not reported) | NA | No | No |
| El-Raheem et al., | 30 | No | NA | NA | No | No |
| Faisal et al., | 323 | Yes | CARMc19_NB = 0.87 (95% CI 0.85-0.89) vs CARMc19_N 0.86 (95% CI 0.84-0.87) | CARMc19_NB = 0.88 vs CARMc19_N = 0.86 | No | Yes |
| Fumagalli et al., | 120 | No | 0.90 (0.87 - 0.93) | NA | No | No |
| Galloway et al., | 244 | No | 0.697 (0.652,0.741) | NA | No | No |
| Gao et al., | 254 | Yes | 0.9621 (95% CI: 0.9464–0.9778) | 0.9760 (0.9613–0.9906), and 0.9246 (0.8763–0.9729) | Yes | No |
| Garibaldi et al., | 131 | No | Not available | Not available | No | No |
| Garrafa et al., | 505 | Yes | Training: 0.98 | Validating and testing: 0.83 and 0.78 respectively | No | Yes |
| Gavelli et al., | NA (consensus) | No | NA | Not reported | No | No |
| Goméz et al., | 33 | No | 0.874 (0.816-0.933) | NA | No | No |
| Gue et al., | 145 | No | 0.793 (95% CI 0.745–0.841) | NA | No | No |
| Hajifathalian et al., | 93 | Yes | 7 days: 0.877 (95%CI 0.831–0.923); 14 days: 0.847 (95%CI 0.806–0.888) | 7 day (0.851 [0.781 to 0.921]); 14 day (0.825 [0.764 to 0.887]) | No | Yes |
| Halalau et al., | Not clear | Yes | Not available | 0.75 (0.71 – 0.78) | No | No |
| Halasz et al., | 293 | Yes | Internal validation: 0.78(95% CI 0.74-0.84, Brier-score 0.19) | External validation: 0.79 (95% CI 0.68-0.89, Brier-score 0.16) | No | No |
| Hohl et al., | 618 | Yes | 0.92 (95% confidence intervals [CI] 0·91–0·93) | 0.92 (95%CI 0·89–0·93) | No | No |
| Hou et al., | 82 | No | 0.88 | Not clear | No | No |
| Hu et al., | 68 | Yes | 0.895 (95% CI not reported) | 0.881 (95% CI not reported) | No | No |
| Incerti et al., | 2163 | No | 0.8822 | 0.8741 | No | No |
| Jamshidi et al., | 2440 (10.27%) | No | 0.74-0.80 | 0.635-0.79 | No | No |
| Kapoor et al., | 33 | NA | No | Illustrated in Figure 2. | No | No |
| Kazemi et al., | 11 | No | 0.73 (95% CI not reported) | NA | No | No |
| Kim et al., | 7 | No | Not reported | NA | No | No |
| Knight et al., | 11426 | Yes | 0.786 (0.781 - 0.790) | 0.767 (0.760 - 0.773) | No | Yes |
| Ko et al., | 212 (58.7%) | Yes | Not reported | Not reported | No | No |
| Le Lannou et al., | 349 | No | AUC-ROC: 0.730, 95% CI: 0.700 -0.760 | NA | No | No |
| Levy et al., | Not clear | Yes | 0.86 (95% CI not reported) | 0.82 (95% CI not reported) | No | No |
| Li et al., | 142 | No | 0.852 | 0.844 | Yes | No |
| Liang et al., | 51 (3.2%) | Yes | 0.88 (0.85 - 0.91) | 0.88 (0.84 - 0.93) | No | No |
| Lin et al., | Not clear | Yes | ANN: 0.96 and CNN: 0.91, using normalized training data | ANN: 0.80 and CNN: 0.73, using normalized testing data | No | No |
| Lu et al., | 39 | No | Not reported | NA | No | No |
| McRae et al., | 753 | Yes | No | No | No | Yes |
| Mei et al., | 105 | Yes | 0.912 (95% CI 0.878-0.947) | VC1 = 0.928 (95% CI 0.884-0.971) and VC2 = 0.883 (0.815-0.952) | No | No |
| Momeni-Boroujeni et al., | 211 | No | No | NA | No | No |
| Monterde et al., | Not clear | No | The AUC for prediction of critical illness was 0.641 (95% CI 0.624-0.660) for the Charlson index, 0.665 (0.645-0.681) for the Elixhauser index, and 0.787 (0.773-0.801) for Queralt DxS. | NA | No | No |
| Naseem et al., | Not clear | No | 869 | No | No | No |
| Nicholson et al., | Not reported | Yes | 0.87 (0.83 – 0.91) | 0.80 (0.75 – 0.85) | No | No |
| Núñez-Gil et al., | 311 | No | 0.88 (0.85 – 0.91) | NA | No | Yes |
| Sarkar et al., | 37 | No | 0.97 (95% CI not reported) | NA | No | No |
| Schlauch et al., | 7327 (15.6%) | No | The RTRM predicted overall mortality as well as mortality 1, 3, and 7 days in advance with an area under the receiver operating characteristic curve (AUCROC) of 0.905, 0.911, 0.905, and 0.901 respectively |  | No | Yes |
| Shang et al., | 49 | Yes | 0.919 (95% CI 0.870-0.970) | 0.938 (95% CI 0.902-0.973) | No | No |
| Soto-Mota et al., | 200 (50%) | No | NA | Provided by different cut-offs, ranging from 0.61 to 0.90 (95% ranges from 0.59 to 0.93), with best AUC for 25 points (0.90 [95% CI 0.87-0.93]) | No | No |
| Sottile et al., | 717 | No | 0.94 | 0.9 | Yes | No |
| Sourij et al., | 58 | No | 0.889 (0.837 - 0.941) | NA | No | No |
| Stachel at al., | 452 (while 776 died in the training, validation and test sets) | No | 0.83 | 0.97 | No | Yes |
| Turrini et al., | 98 | No | Only for some variables, as: age distribution and P/F ratio at presentation showed an optimal cut-point at 69 years (AUC 0.71,) and at 233 (AUC 0.73), respectively. Inflammation, LDH and CRP showed an optimal cut-point at 395 U/L (AUC 0.77) and at 124 mg/L (AUC 0.70) respectively. Between the 29 patients tested, IL6 values showed an optimal cut-point at 3484 pg/mL (AUC 0.862). | NA | No | No |
| Vaid et al., | 510 | No | 0.693 to 0.805 | No | Yes | Yes |
| Vaid et al., | Not clear | Yes | AUC-ROC of 0.89 at 3 days, 0.85 at 5 and 7 days, and 0.84 at 10 days | AUC-ROC of 0.88 at 3 days, 0.86 at 5 days, 0.86 at 7 days, and 0.84 at 10 days | Yes | Yes |
| van Dam et al., | 167 | Yes | 0.84 | The model yielded an AUC of 0.77 (95% CI 0.73 to 0.81) for 30-day mortality and an AUC of 0.72 (95% CI 0.68 to 0.76) for a composite of 30-day mortality and/or admission to ICU | No | Yes |
| Vela et al., | 19114 | No | 0.96 (0.96 – 0.96) | 0.96 (0.96 – 0.96) | No | No |
| Wang et al., | 24 | Yes | 0.871 (based on its optimal cut-off value = 85) | Not available (link for supplemental material does not work) | No | No |
| Weng et al., | 21 | Yes | 0.921 (0.835-0.968) | 0.975 (0.947-1.0) | No | No |
| Williams et al., | 11407 | Yes | 0.896 (95% CI 0.72 - 0.90) | CUIMC database 0.820 (95% CI 0.796-0.840); HIRA database 0.898 (95% CI 0.857-0.940); SIDIAP 0.895 (95% CI 0.881-0.910); VA 0.717 (0.642-0.791) | No | Yes |
| Wollenstein-Betech et al., | Not clear | No | 0.69 | No | Yes | No |
| Wu et al., | Not clear | Yes | Model 1 with AUC of 0.74 (95% CI 0.69–0.79) on the training dataset, Model 2 with the clinical features of age, hospital employment, body temperature and the time of  onset yielded an AUC of 0.78 (95% CI 0.73–0.83) on the training dataset and Model 3 was based on age and lesion range score on CT and had an AUC of 0.75  (95% CI 0.70–0.80) on the training dataset. | Model 1 with AUC of 0.83 (95% CI 0.72–0.94) on the validation dataset, Model 2 with AUC of 0.74 (95% CI, 0.59–0.89) and Model 3 had AUC of 0.83 (95% CI, 0.72–0.94). | No | No |
| Xie et al., | 155 | Yes | 0.880 (95% CI not reported) | 0.980 (0.958-1.00) | No | Yes |
| Yadaw et al., | 313 (8.15%) | Yes | 0.91 (95% CI not provided) | 0.91 (95% CI not provided) | No | Yes |
| Yan et al., | 174 | Yes | 0.978 (IC 95% not provided) | 0.951 (CI 95% not provided) | Yes | No |
| Yoo et al., | Not reported | Yes | Not reported, as AUC was used to define the variables for the score. | At admission 0.81; maximum through admission 0.91; mean through admission 0.92 | No | No |
| Yu et al., | 506 | No | 0.90 | 0.85 | No | No |
| Zhang et al., | 96 | Yes | 0.886 (95% CI 0.873–0.899) | 0.879 (95% CI, 0.856–0.900) and 0.839 (95% CI [0.798–0.880) for each one of the hospitals | No | No |
| Zhang et al., | 33 (4.3%) | Yes | DCS: 0.79; DCS: 0.89; DL: 0.91 (95% CI not reported) | DL: 0.74 (95% CI not reported) | No | Yes |
| Zhao et al., | 82 | No | 0.87 ([95% CI, 0.83–0.92], p<0.001) for mortality | 0.82 ([95% CI, 0.73–0.92], p<0.001) | No | Yes |
| Zhou et al., | 54 | No | No | No | No | No |
| Zhou et al., | 51 | No | 0.955 (95% CI not provided) | NA | No | No |
| Zhu et al., | 39 | No | 0.968 (95% CI = 0.87–1.0) | 0.954 (95% CI = 0.80–0.99) | No | No |

ARDS: acute respiratory distress syndrome; AST: aspartate transaminase; AUC: area under the curve; BMI: body mass index; BUN: blood urea nitrogen; CCEDRRN: canadian covid-19 emergency department rapid response network; CI: confidence interval; CKD: chronic kidney disease; COPD: chronic obstructive pulmonary disease; CPR: C-reactive protein; CT: computed tomography; DLN: deep learning networks; DM: diabetes mellitus; ED: emergency department; EH: emergency hospital; ER: emergency room; FiO2: fraction of inspired oxygen; GFR: glomerular filtration rate; GP: general practice; ICU: intensive care unit; IHD: ischemic heart disease; IL-6: interleukin 6; INR: international normalized ratio; LASSO: least absolute shrinkage and selection operator logistic regression; LDH: lactate dehydrogenase; MAP: mean arterial pressure; MICE: Multivariate Imputation by Chained Equations; NA: not applicable; NAAT: nucleic acid amplification test; NEWS2: national early warning score; NLR: neutrophil lymphocyte ratio; OP: outpatient; PLS: partial least squares; RDW: red blood cell distribution width; RF: Random Forest; RT-PCR: reverse transcription polymerase chain reaction; SF ratio: SpO_2_/FiO_2_ ratio; SVM: support-vector machine; Trop I: troponin I; XGBoost: eXtreme Gradient Boosting; WBC: white blood cell; WHO: World Health Organization.
