## Supplementary material for "Effectiveness, Explainability and Reliability of Machine Meta-Learning Methods for Predicting Mortality in Patients with COVID-19: Results of the Brazilian COVID-19 Registry": Table 2

**Table S2. Potential predictors included for the development of the models**

**Variables**

**Demographics characteristics**

Sex at birth

Age (years)

**Comorbidities**

Hypertension

Coronary artery disease

Heart failure

Atrial fibrillation or flutter

Stroke

Chagas disease

Rheumatic heart disease

Other cardiovascular disease

No relevant cardiovascular disease

Asthma

COPD

Pulmonary fibrosis

Diabetes mellitus

Obesity (BMI>30 kg/m^2^)

Cirrhosis

Psychiatry disease

Chronic kidney disease

Rheumatologic disease

HIV infection

Cancer

Previous organ transplantation

Immunosuppressive condition

Another relevant health condition

Number of comorbidities

Number of cardiovascular comorbidities

**Lifestyle habits**

Illegal drug use

Alcoholism

Current smoker

Ex-smoker

**Clinical characteristics**

Time from symptom onset

Respiratory rate (irpm)

Heart rate (bpm)

Systolic blood pressure (mmHg)

Diastolic blood pressure (mmHg)

Inotrope use

Glasgow coma score

SF ratio

**Laboratory**

C reactive protein (mg/L)

Hemoglobin (g/L)

Leucocytes (10^9^/L)

Neutrophils (10^9^/L)

Lymphocytes (10^9^/L)

Neutrophils-to-lymphocytes ratio

Platelet count (10^9^/L)

Creatinine (mg/dL)

Urea (mg/dL)

Lactate (mmol/L)

Sodium (mmol/L)

Bicarbonate (mEq/L)

pH

pO_2_ (mmHg)

pCO_2_ (mmHg)

D-dimer

BMI: body mass index; COPD: chronic obstructive pulmonary disease; HIV: human immunodeficiency viruses; pO_2_: partial pressure of oxygen; PCO_2_: partial pressure of carbon dioxide; SF ratio: SpO_2_/FiO_2_ ratio
